# Gastroenterological and Hepatic Manifestations of Patients with COVID-19, Prevalence, Mortality by Country, and Intensive Care Admission Rate: Systematic Review and Meta-analysis

**DOI:** 10.1101/2020.10.29.20207167

**Authors:** Mohammad Shehab, Fatema Alrashed, Sameera Shuaibi, Dhuha Alajmi, Alan Barkun

## Abstract

**Background & Aims:** Patient infected with the SARS-COV2 usually report fever and respiratory symptoms. However, multiple gastrointestinal (GI) manifestations such as diarrhea and abdominal pain has been described. The aim of this study was to evaluate the prevalence of GI, liver function test (LFT) abnormalities, and mortality of COVID-19 patients.

**Methods:** We performed a systematic review and meta-analysis of published studies that included cohort of patients infected with SARS-COV2 from December 1^st^, 2019 to July 1^st^, 2020. We collected data from the cohort of patients with COVID-19 by conducting a literature search using PubMed, Embase, Scopus, and Cochrane according to the preferred reporting Items for Systematic Reviews and MetaAnalyses (PRISMA) guidelines. We analyzed pooled data on the prevalence of overall and individual gastrointestinal symptoms, LFTs abnormalities and performed subanalyses to investigate the relationship between gastrointestinal symptoms, geographic location, fatality, and ICU admission.

**Results:** The available data of 17,802 positive patients for SARS-COV2 from 120 studies were included in our analysis. The most frequent manifestations were diarrhea (13.3%, 95% CI 12-16), nausea (9.1%, 95% CI 9-13) and elevated LFTs (23.7%, 95% CI 21- 27). The overall and GI fatality were 7.2% (95% CI 6 -10), and 1% (95% CI 1- 4) respectively. Subgroup analysis showed non statistically significant associations between GI symptoms/LFTs abnormalities and ICU admissions (OR=3.41, 95% CI 0.87 – 13.4). The GI mortality rate was 0.58% in China and 3.5% in the United States (95% CI 2 - 5).

**Conclusion:** Digestive symptoms and LFTs abnormalities are common in COVID-19 patients. Our subanalysis shows that the presence of gastrointestinal and liver manifestations does not appear to affect mortality, or ICU admission rate. However, the mortality rate was higher in the United States compared to China.

## Introduction

In December 2019 China was struck with a new strain of Coronavirus, Novel Coronavirus (2019 nCov). Within a short period of time, it soon spread to a full pandemic.

It was first noticed by the innumerable cases of pneumonia that suddenly surged amongst local inhabitants in the Province of Wuhan. Soon, the virus was detected through sequencing leading to officially being renamed severe acute respiratory syndrome coronavirus 2 (SARS-COV2) by the International Committee on Taxonomy of Viruses [1-2]. The disease caused by Coronavirus (SARS-COV2) was allocated the title of COVID-19 or ‘Coronavirus disease’ [3]. Coronaviruses in general are single stranded RNA viruses falling under the family of Coronaviridae which also include MERS (MERS Cov) and SARS (SARS Cov)[4]. By the end of May, 5,796, 257 cases of COVID-19 have officially been confirmed worldwide and as of July the number of confirmed cases passed the 12 million mark [5].

It has been established that the transmission of Coronavirus (SARS-COV2) occurs from person to person through upper airway tract (droplet infection) or direct contact [6]. The virus can also be detected in saliva, urine, gastrointestinal tract and possibly through airborne spread [7,8]. The spectrum of symptoms attributable to SARS-COV2 include fever, cough, myalgia, fatigue, and to a lesser extent headache. Patients may also be asymptomatic [9-11]. Diarrhea, nausea and vomiting, as well as liver involvement have all been reported in the literature [12,13]. In fact, gastrointestinal involvement is plausible given that angiotensin converting enzyme 2 (ACE2), the major human cellular receptor for the SARS-COV2, is expressed in the gastrointestinal tract as well as liver cells [14]. We thus conducted a systematic review of published gastrointestinal and liver symptoms associated with COVID-19 on the basis of disease severity, age group, and geographical region.

## Methods

### Search strategy

A systematic review was conducted using Pubmed, Scopus, Cochrane, and Embase databases. Medical literature searches for human studies were performed from December 1^st^, 2019 up to July 1^st^, 2020. The key terms used for the literature search were ((“COVID-19” OR “COVID 2019” OR “severe acute respiratory syndrome coronavirus 2” OR “severe acute respiratory syndrome coronavirus 2” OR “2019 nCoV” OR “SARS-COV2” OR “2019nCoV” OR (“severe acute respiratory syndrome coronavirus 2” OR “SARS-COV2” AND GASTROINTESTINAL AND (MANIFESTATIONS OR CLINICAL CHARACHTERISTICS) OR (“gastrointestinal tract” OR (“gastrointestinal’ AND “tract”) OR “gastrointestinal tract” OR (“gi” AND “tract”) OR (“fatality” or “Mortality”). In addition, a manual search of all review articles, editorials and retrieved original studies was also performed. All procedures used in this meta-analysis were consistent with the Preferred Reporting Items for Systematic Reviews and Meta analyses (PRISMA) guidelines and prespecified protocol, which described our method and analysis before data collection was initiated (see supplementary PRISMA check list).

### Selection criteria and data extraction

Data were independently abstracted based on our protocol by two investigators (MS and FA) and any discrepancies between the two authors were resolved through discussion. Inclusion and exclusion criteria were defined prior to the literature search. The inclusion criteria were (1) study type: case reports/case series (including chart reviews), prospective/retrospective cohort studies, case control studies, cross sectional studies and randomized controlled trials; (2) patients population: Adults patients with COVID-19; inpatient or outpatient setting and (3) Outcome measured: At least one GI manifestation reported and LFT abnormality, defined as any value above the normal upper limit.

Exclusion criteria were (1) Review, opinion, abstracts from conferences, editorials, commentary articles, review articles and meta analyses; (2) studies without data for retrieval; (3) duplicate studies; (4) Asymptomatic patients with COVID-19: (5) studies that did not report gastrointestinal symptoms.

Data extraction was performed using Microsoft excel. The following data were extracted:

1. Study: author, journal, date, country, number of patients, and study type.
2. Patients characteristics: mean age, ethnicity, gender, and comorbidities.
3. Overall fatality rate and fatality rate by country.
4. Number of patients admitted to the ICU.
5. Gastrointestinal symptoms proportion: abdominal pain, diarrhea, nausea, anorexia, loss of taste, elevated liver enzymes or other nonspecified gastrointestinal symptoms.

### Risk of bias and certainty of evidence

The Methodical Index for Nonrandomized Studies (MINORS) [15] was used to assess bias risk (see table 2). In addition, risk of bias was assessed based on 4 domains: selection, ascertainment, causality, and reporting. An overall judgment of risk of bias was made based on factors deemed to be most critical for the systematic review (selection criteria, ascertainment of outcome, and followup duration).

### Statistical Analysis

Our primary analysis focused on assessing the weighted pooled prevalence of GI symptoms/LFT abnormalities in patients with COVID-19 infection, occurring any time during the course of illness. We also conducted subanalyses that looked at the association between GI symptoms/LFT abnormalities and mortality as well as ICU admission. Categorical variables were described as count (%). Continuous variables were described using mean (SD) if they are normally distributed, median (IQR) if they are not. We pooled the single arm event rates using a random effects method and we measured heterogeneity within our studies using the I^2^ statistic. Subanalyses were described and tested using odd ratios and 95% confidence interval to determine statistical significance. Stata 16 was used to calculate odd ratios and their respective 95% CI and create Forest and Box plots.

### Sensitivity Analysis

To examine the effect of the quality of studies on our results, we performed a sensitivity analysis on the prevalence of GI symptoms and LFTs abnormalities by excluding low qualities studies. These include case reports, case series and case control studies. We also assessed selected outcomes comparing patients with versus without GI/LFTs abnormalities.

## RESULTS

### Research selection and quality assessment

Overall, 120 studies (supplementary table. S1) from 2355 potentially relevant citations were included in the analysis (Figure 1). Most of the included studies were single arm only, very few studies included comparator groups. Furthermore, outcome assessors in all 120 studies were not blinded. Both inpatients and outpatients studies were included. The risk of evidence imprecision was rated as very serious, given that the included studies were all observational studies. Overall, all included studies were rated as having very serious risk of bias because they lacked a control group and had a high risk of confounding and selection bias.

**Figure 1:**
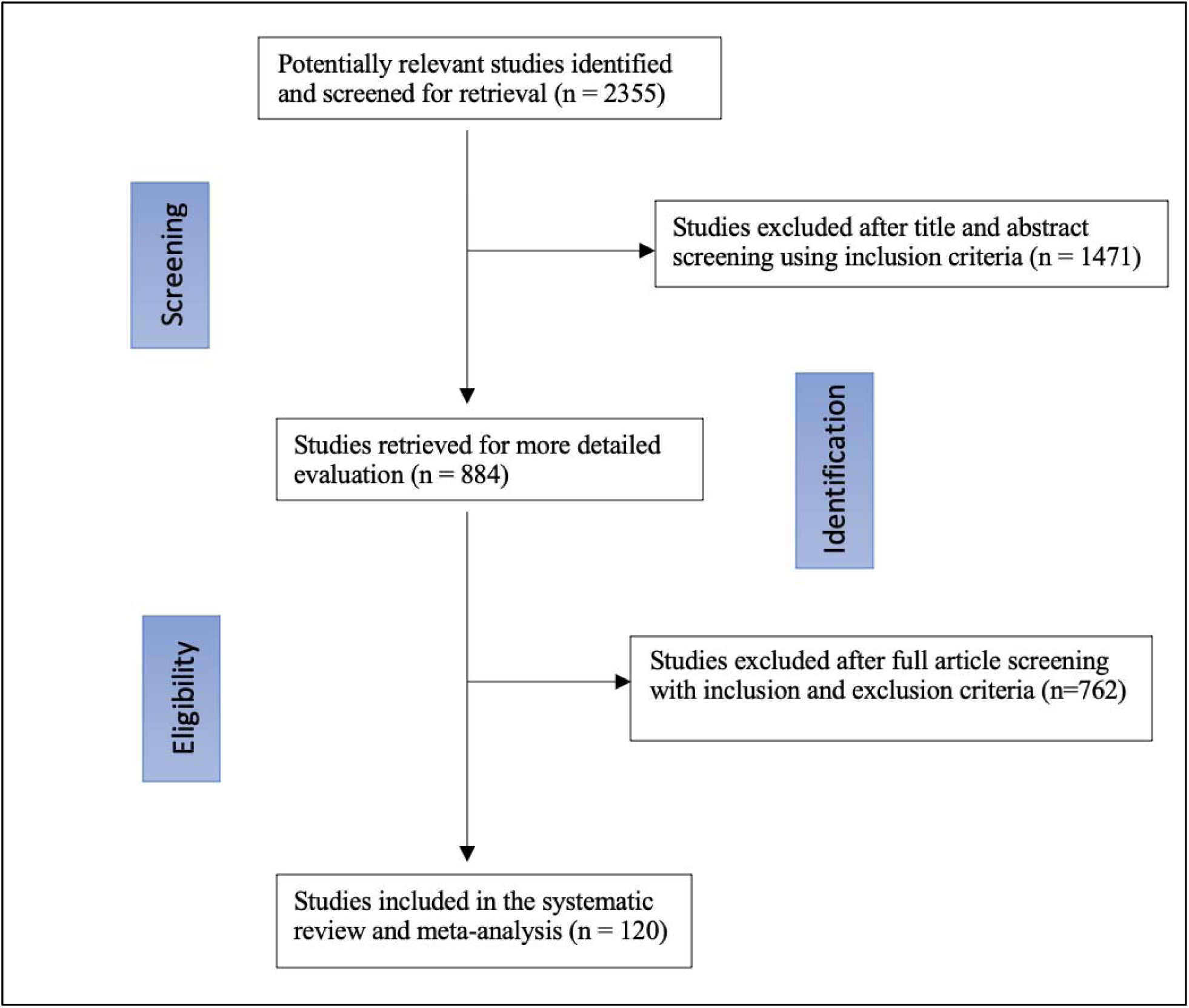
Flow diagram for study selection

### Clinical data

This systematic review included 120 studies [16-136] with a total of 17,801 patients that tested positive for SARS-COV2 and were included in the analysis. The mean patient age was 52.6 (±14, 95% CI 48-57.3) and 47.6% of the patients were males. Most patients had several comorbidities, most common being hypertension (23.4%, 95% CI 21-27), diabetes mellites (13.4%, 95% CI, 12-15), and cardiovascular diseases (12.4%. 95% CI, 10-14). GI symptoms included nausea, vomiting, and diarrhea. Heterogeneity statistic I2 is 94.5%, which signifies a significant heterogeneity among our studies. The most common GI/liver manifestations were elevated liver enzymes and diarrhea (supplementary figure.1). Specifically, GI manifestations of patients infected with SARS-COV2 are diarrhea 13.3% (95% CI 12 - 16), nausea (9.1%, 95% CI 9 - 13) anorexia or loss of appetite (8.3%, 95% CI 8 - 11), vomiting (6.4%, 95% CI 5 - 8), abdominal pain (3%, 95% CI, 3-6), loss of taste (1.3%, 95% CI 1 - 4), and elevated LFTs (23.7%, 95% CI 21 - 27).

### Sensitivity Analysis

The sensitivity analysis included 25 studies (supplementary: table S4). The results did not differ from our main analysis. Among the GI manifestations experienced by COVID-19 patients, diarrhea (13.1%, 95% CI, 12-14), was still the most common symptom, followed by nausea (9.2%, 95% CI 8-11). The percentage of patients experiencing LFTs abnormalities was 24% (95% CI 21 - 26).

### Fatality rates and Geographic Variation

A total of 78 studies reported the incidence of mortality. The overall fatality rate of patients was 7% (95% CI 6 -10), (supplementary: table S5). The subgroup analysis included three studies [19-21] that directly compared mortality rate in patients with and without GI symptoms. In this analysis the number of patients who experienced gastrointestinal symptoms/liver test anomalies and those who did not were 227 and 326, respectively. The results showed that patients with GI manifestations//LFT abnormalities were no more likely to die compared to those who did not with a pooled odd of patients was not statistically significant 1.07 (95% CI 0.58 - 1.97) (figure 3).

**Figure 2:**
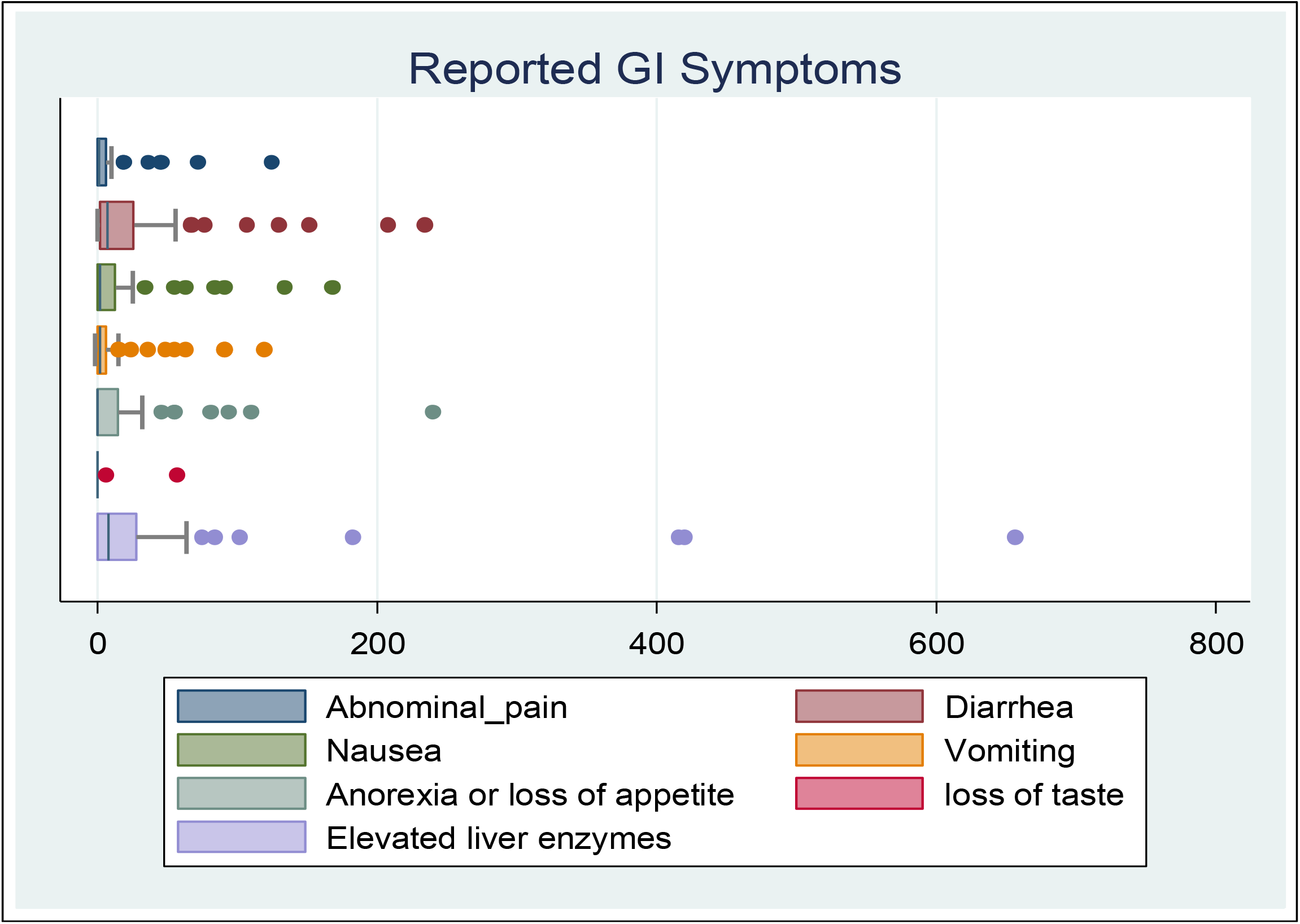
Box plots representing the range and outliers of the gastrointestinal and liver manifestations in patients with COVID-19.

**Figure 3:**
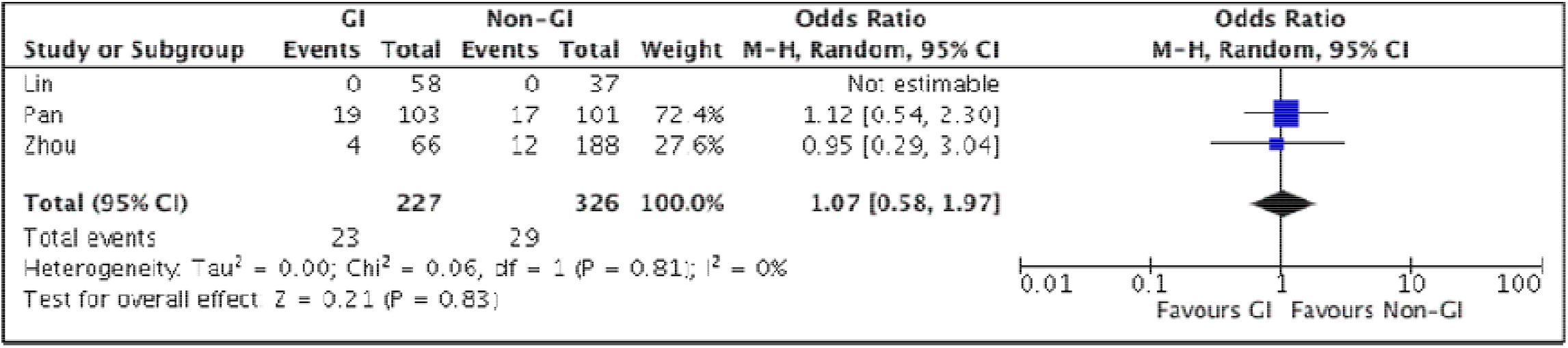
Forest Plot for mortality in patients with COVID-19, showing no significant difference in the pooled odds of patients with gastrointestinal symptoms and those without gastrointestinal adverse events.

Moreover, out of the 78 studies, a total of 25 studies reported mortality in patients with GI/Liver manifestations (see figure 4). A subanalysis of fatality rate in patients with GI symptoms based on their location showed that 27 out 4660 patients (0.58%) in China died (95% CI 0.5, 1.6), whereas 16 out 449 patients (3.5%) in the United States died (95% CI 2 - 5). In addition, 3 studies from Taiwan, Korea, and Japan reported zero fatality (table.1).

**Table. 1.** Mortality by geographic location

| Study | Total no. of patients | Mortality in patients with GI symptoms | country |
| --- | --- | --- | --- |
| Huang C et. al. | 41 | 0 | China |
| Fan H et. al. | 101 | 9 | China |
| Han C et. al. | 206 | 0 | China |
| Huang WH et. al. | 2 | 0 | China |
| Jin X et. al. | 651 | 0 | China |
| Kuang Y et. al. | 944 | 0 | China |
| Lin L et. al. | 95 | 0 | China |
| Luo S et. al. | 1141 | 7 | China |
| Pan F et. al. | 21 | 0 | China |
| Shu L et. al. | 545 | 0 | China |
| Song F et. al. | 51 | 0 | China |
| Wan Y et. al. | 230 | 4 | China |
| Wei X-S et. al. | 84 | 0 | China |
| Wu Y et. al. | 74 | 0 | China |
| Xia X et. al. | 10 | 0 | China |
| Zhao D et. al. | 19 | 0 | China |
| Zhou F et. al. | 191 | 2 | China |
| Zhou Z et. al. | 254 | 5 | China |
| **Total China** | **4660** | **27** | **0.58%** |
| Cholankeril G et. al. | 116 | 0 | US |
| Kujawski S et. al. | 12 | 0 | US |
| Redd W et. al. | 318 | 16 | US |
| Siegel et. al. | 3 | 0 | US |
| **Total USA** | **449** | **16** | **3.5%** |
| Hsih W-H et. al. | 2 | 0 | Taiwan |
| Tabata S et. al. | 104 | 0 | Japan |
| Kim ES et. al. | 28 | 0 | Korea |

**Figure 4:**
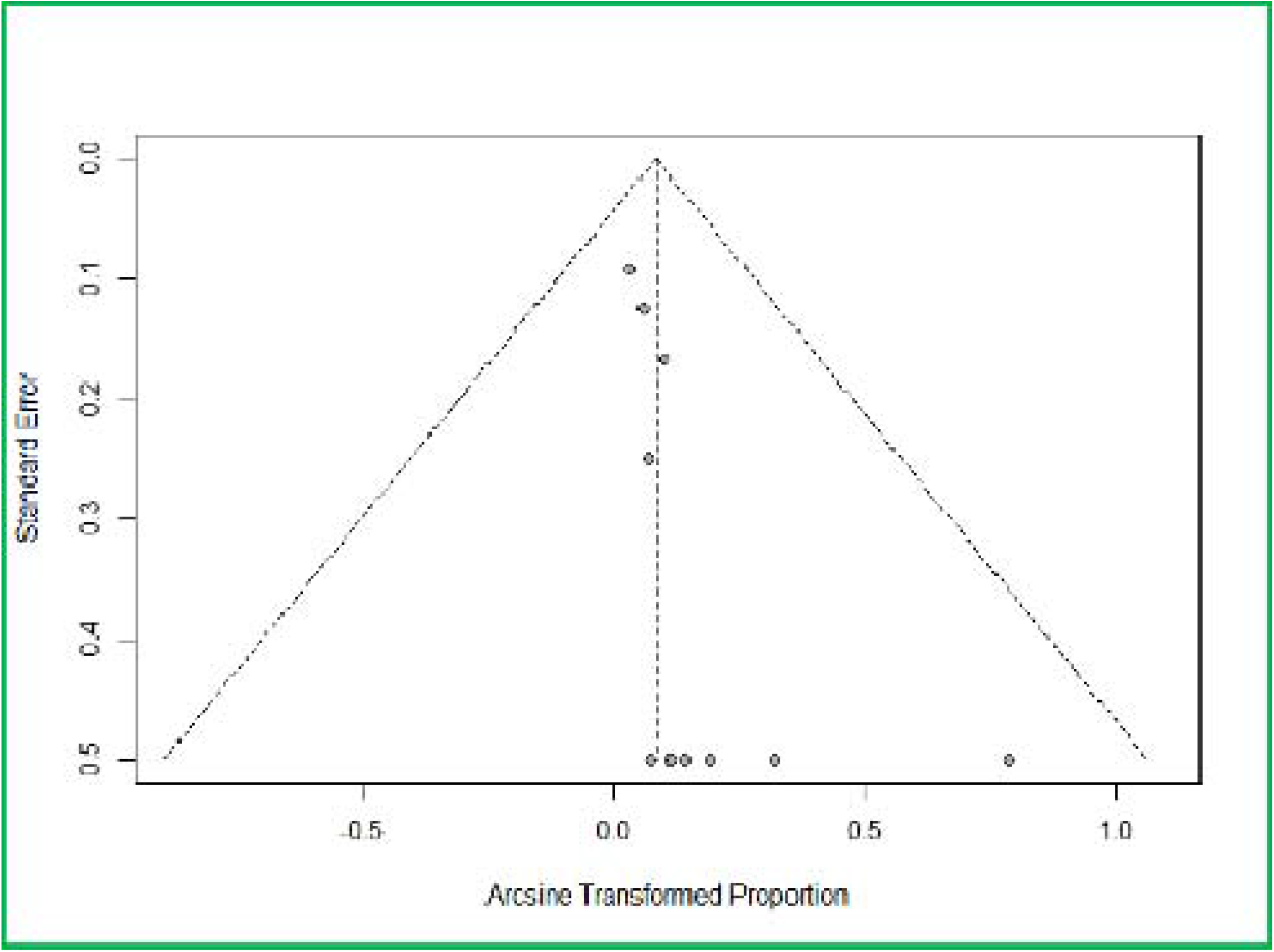
Funnel plot of the 25 studies that reported mortality of patients with GI symptoms

### ICU admission rate

Only two studies [20,21] reported differences in ICU admissions amongst patients manifesting GI symptoms/LFT abnormalities and patients without. The total number of patients with gastrointestinal problems or LFT abnormalities who were admitted in the ICU were 23 and the number of patients who did not experience gastrointestinal problems or LFT abnormalities and were admitted to the ICU were 156. No statistically significant difference in ICU admission rate was noted between those who experienced GI/LFTs abnormalities and those who did not. The pooled proportion was 3.41 (95% CI, 0.87,13.4). (Figure. 5).

**Figure 5:**
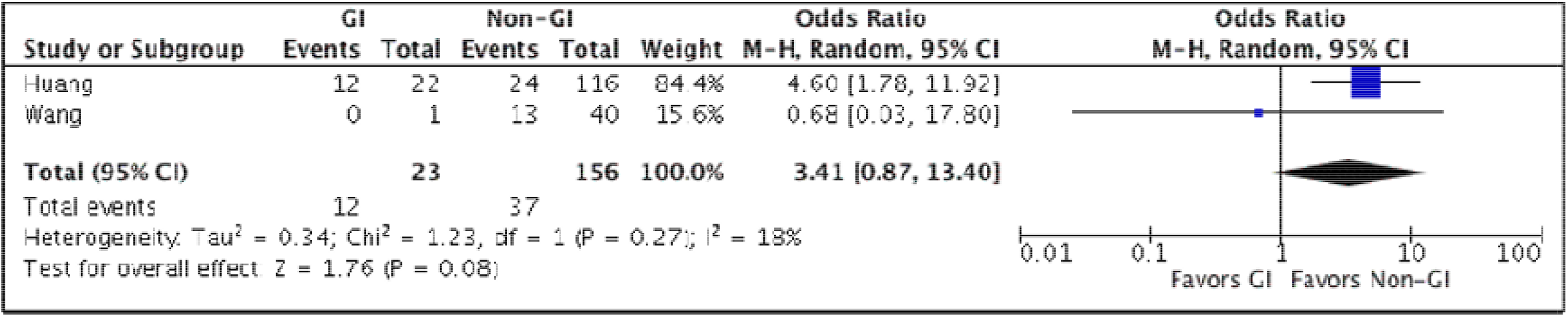
Odds ratio of ICU admissions in patients with COVID-19amongst patients with and without gastrointestinal symptoms / LFT abnormalities.

## Discussion

This meta-analysis of 17,801 COVID-19 patients found that gastrointestinal symptoms are common in patients infected with SARS-COV2. Our study has several strengths. This is one of the more recent meta-analyses that summarizes the rapidly emerging and sometimes confusing literature on COVID-19 on the prevalence of overall and individual gastrointestinal manifestations. The large patient population and comprehensive inclusion of 120 studies allow a more precise estimation of the prevalence of gastrointestinal symptoms and liver test anomalies associated with COVID-19. Subgroup analysis found that no association between the presence of gastrointestinal symptoms/LFT anomalies and mortality; but a trend towards their absence and ICU admission was noted.

Gastrointestinal symptoms including abdominal pain, diarrhea, nausea, vomiting, loss of appetite, loss of taste and elevated liver enzyme are among the presenting symptoms or laboratory abnormalities of SARS-COV2 infection. Diarrhea was the most common gastrointestinal symptom; this is particularly important because previous studies have shown that patients with diarrhea on presentation have a higher stool RNA positivity and viral load than those without [137,138]. One study showed that 44 of 153 patients with COVID-19 tested positive for the virus in the stools [139]. Another study by Xiao et al. showed that among 73 hospitalized COVID-19 patients in China, 39 (53.42%) tested positive for SARS-COV2 RNA in stools [140]. In addition, a report of a COVID-19 patient with positive fecal but negative pharyngeal and sputum viral tests has been described [140]. This may imply that fecal oral route is a possible route of SARS-COV2 transmission.

The possibility of fecal oral transmission of SARS-COV2 emphasizes the importance of frequent and proper hand hygiene. This is important in every clinical setting, but especially in low resource areas with poor sanitation. Intuitively, proper handling of the excreta of COVID-19 patients should still be strongly enforced, and sewage from hospitals should also be properly disinfected. The presence of the virus in the digestive tract also raises the concerns of COVID-19 infection in patients with established gastrointestinal conditions as well as potential fecal microbiota transplant donors. Nevertheless, the unknown effect of COVID-19 on patients with preexisting gastrointestinal diseases and its influence on treatment and outcome is a cause for concern [140]. These implications warrant further investigation. The American Gastroenterological Association (AGA) and joint society recommend the use of enhanced personal protective equipment, including the use of N95 (or N99) masks instead of surgical masks, for health care workers performing upper or lower GI procedures regardless of COVID19 status [141].

It is believed that the prevalence of gastrointestinal symptoms is underestimated because the majority of studies only reported gastrointestinal symptoms on the day of admission but not throughout the disease course [142]. Furthermore, many earlier studies did not report on other gastrointestinal symptoms except for diarrhea [141,143]. Based on these findings, clinicians must be aware that digestive symptoms, such as diarrhea, may be a presenting feature of COVID-19 that can arise before respiratory symptoms, and on rare occasions may be the only presenting manifestation of COVID-19.

The analysis also found that elevated liver enzymes are a common laboratory marker of COVID-19 patients. Huang et al. showed that AST elevation was observed in 8 (62%) of 13 patients in the intensive care unit compared with only 7 (25%) of 28 patients who did not require care in an ICU [140]. On the other hand, one study from China showed that COVID-19 patients with underlying chronic hepatitis B infection did not have higher disease severity compared to the overall population [143]. Unfortunately, most studies report liver enzymes as the mean/median value of the entire cohort without cutoff values for a given institution rather than as a proportion of patients with elevated values. One aspect that remains to be determined is the impact of COVID-19 in patients with preexisting chronic liver diseases, such as viral hepatitis, and fatty liver disease.

The pooled analysis showed that the overall fatality rate was 11.2%. While the finding is not statistically significant, any possible true difference in mortality may be worth further investigation among better defined COVID-19 patient subgroups with GI/LFT anomalies because one study showed that prevalence of severe disease was more common in patients who had gastrointestinal symptoms than those who did not [141]. Furthermore, a study by Wang et al found that abdominal pain was more frequent in patients who required ICU care than those who did not [142]. Although our finding was not statistically significant, the subgroup sub analysis showed that patients who did not have gastrointestinal manifestations were more likely to be admitted to the ICU. This possible finding also requires additional data.

### Limitations

Most of the studies we base our analyses on are observational, single arm cohorts. The lack of control groups and comparison arms can lead to bias due to confounding. Also, our subanalyses might have been affected by small sample sizes. Additionally, regarding fatality rate among COVID-19 patients, most of the studies did not differentiate between the GI symptoms and LFTs abnormalities when performing head to head comparison.

## Conclusion

In this meta-analysis, we summarize the recent reports of digestive symptoms/LFT anomalies among patients infected with SARS-COV2. Gastrointestinal symptoms are commonly observed in patients with COVID-19, therefore, clinicians should be aware that diarrhea and nausea can be the only manifestations of COVID-19 patients. Our sub analysis showed that patients infected with SARS-COV2 and exhibiting digestive symptoms had higher mortality rate in the United States compared to China. We also could not find statistically significant association between ICU admission in patients with GI symptoms compared to those without digestive symptoms or hepatic manifestations due to small sample size; however, further investigation is warranted to better assess this possible association.

## Data Availability

all data link availabe

---

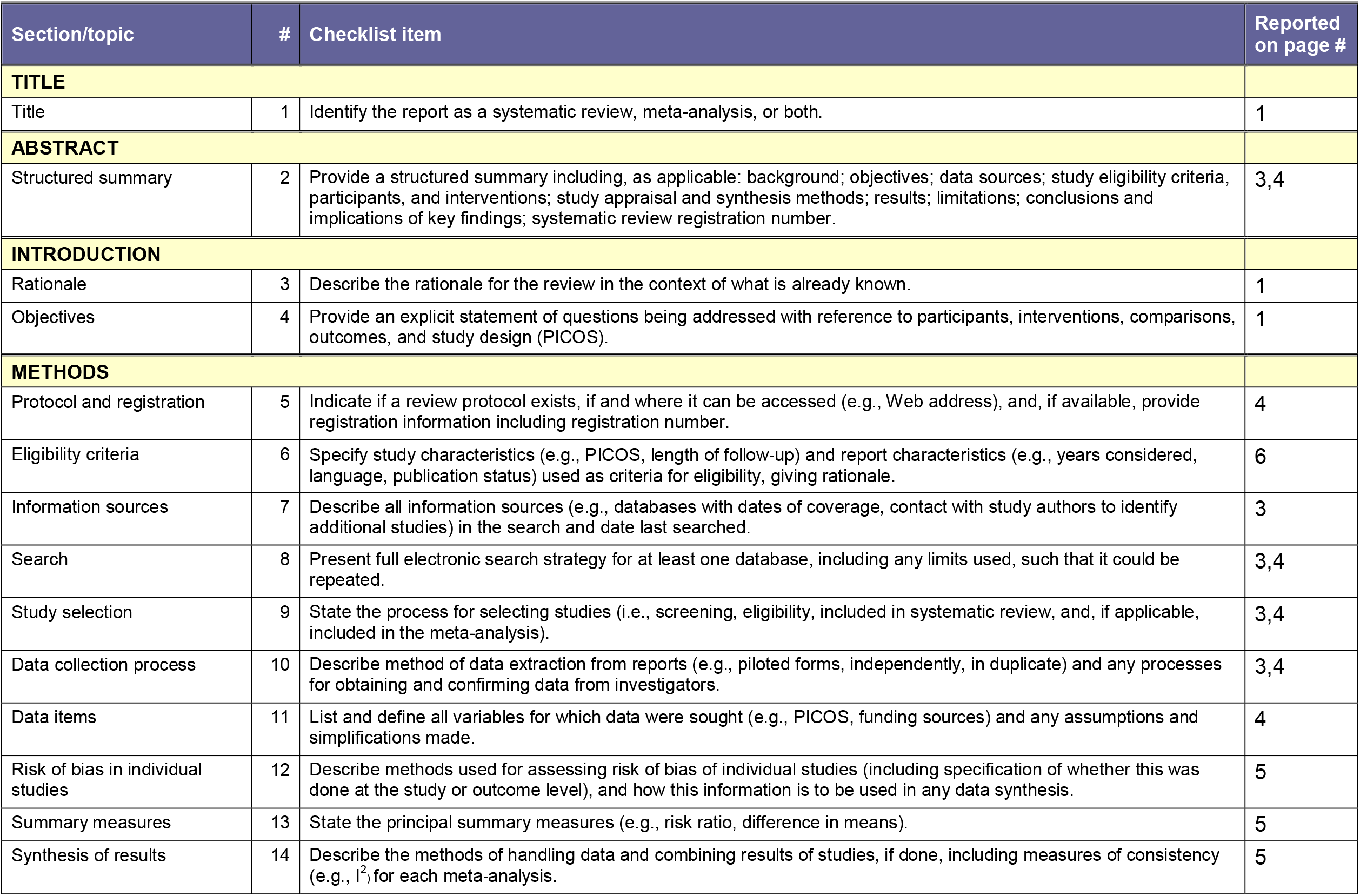
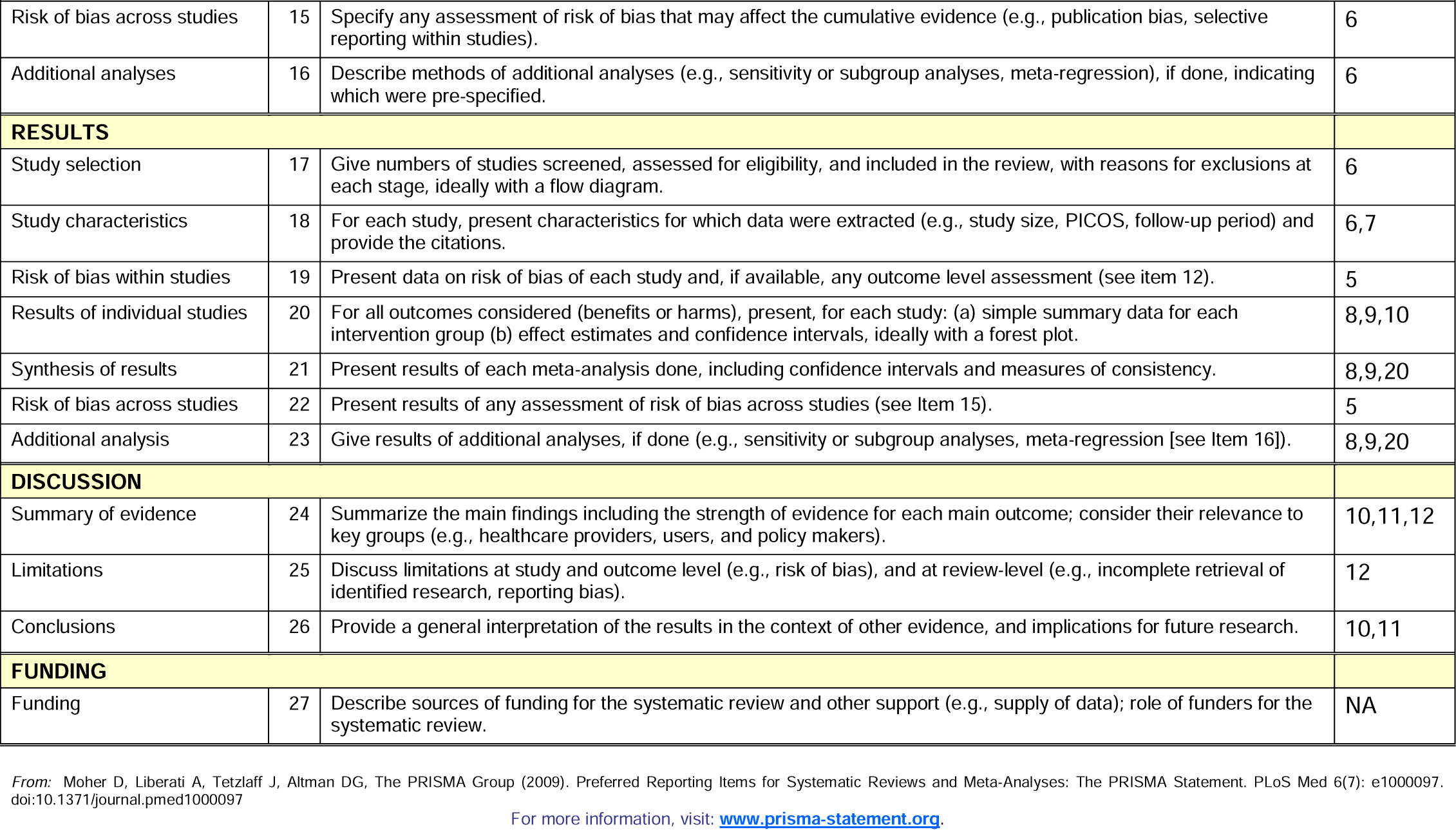

PRISMA 2009 Checklist
| Section/topic | # | Checklist item | Reported on page # |
| --- | --- | --- | --- |
| **TITLE** |  |  |  |
| Title | 1 | Identify the report as a systematic review, meta-analysis, or both. | 1 |
| **ABSTRACT** |  |  |  |
| Structured summary | 2 | Provide a structured summary including, as applicable: background; objectives; data sources; study eligibility criteria, participants, and interventions; study appraisal and synthesis methods; results; limitations; conclusions and implications of key findings; systematic review registration number. | 3,4 |
| **INTRODUCTION** |  |  |  |
| Rationale | 3 | Describe the rationale for the review in the context of what is already known. | 1 |
| Objectives | 4 | Provide an explicit statement of questions being addressed with reference to participants, interventions, comparisons, outcomes, and study design (PICOS). | 1 |
| **METHODS** |  |  |  |
| Protocol and registration | 5 | Indicate if a review protocol exists, if and where it can be accessed (e.g., Web address), and, if available, provide registration information including registration number. | 4 |
| Eligibility criteria | 6 | Specify study characteristics (e.g., PICOS, length of follow-up) and report characteristics (e.g., years considered, language, publication status) used as criteria for eligibility, giving rationale. | 6 |
| Information sources | 7 | Describe all information sources (e.g., databases with dates of coverage, contact with study authors to identify additional studies) in the search and date last searched. | 3 |
| Search | 8 | Present full electronic search strategy for at least one database, including any limits used, such that it could be repeated. | 3,4 |
| Study selection | 9 | State the process for selecting studies (i.e., screening, eligibility, included in systematic review, and, if applicable, included in the meta-analysis). | 3,4 |
| Data collection process | 10 | Describe method of data extraction from reports (e.g., piloted forms, independently, in duplicate) and any processes for obtaining and confirming data from investigators. | 3,4 |
| Data items | 11 | List and define all variables for which data were sought (e.g., PICOS, funding sources) and any assumptions and simplifications made. | 4 |
| Risk of bias in individual studies | 12 | Describe methods used for assessing risk of bias of individual studies (including specification of whether this was done at the study or outcome level), and how this information is to be used in any data synthesis. | 5 |
| Summary measures | 13 | State the principal summary measures (e.g., risk ratio, difference in means). | 5 |
| Synthesis of results | 14 | Describe the methods of handling data and combining results of studies, if done, including measures of consistency (e.g., $I^2$) for each meta-analysis. | 5 |
| Risk of bias across studies | 15 | Specify any assessment of risk of bias that may affect the cumulative evidence (e.g., publication bias, selective reporting within studies). | 6 |
| Additional analyses | 16 | Describe methods of additional analyses (e.g., sensitivity or subgroup analyses, meta-regression), if done, indicating which were pre-specified. | 6 |
| **RESULTS** |  |  |  |
| Study selection | 17 | Give numbers of studies screened, assessed for eligibility, and included in the review, with reasons for exclusions at each stage, ideally with a flow diagram. | 6 |
| Study characteristics | 18 | For each study, present characteristics for which data were extracted (e.g., study size, PICOS, follow-up period) and provide the citations. | 6,7 |
| Risk of bias within studies | 19 | Present data on risk of bias of each study and, if available, any outcome level assessment (see item 12). | 5 |
| Results of individual studies | 20 | For all outcomes considered (benefits or harms), present, for each study: (a) simple summary data for each intervention group (b) effect estimates and confidence intervals, ideally with a forest plot. | 8,9,10 |
| Synthesis of results | 21 | Present results of each meta-analysis done, including confidence intervals and measures of consistency. | 8,9,20 |
| Risk of bias across studies | 22 | Present results of any assessment of risk of bias across studies (see Item 15). | 5 |
| Additional analysis | 23 | Give results of additional analyses, if done (e.g., sensitivity or subgroup analyses, meta-regression [see Item 16]). | 8,9,20 |
| **DISCUSSION** |  |  |  |
| Summary of evidence | 24 | Summarize the main findings including the strength of evidence for each main outcome; consider their relevance to key groups (e.g., healthcare providers, users, and policy makers). | 10,11,12 |
| Limitations | 25 | Discuss limitations at study and outcome level (e.g., risk of bias), and at review-level (e.g., incomplete retrieval of identified research, reporting bias). | 12 |
| Conclusions | 26 | Provide a general interpretation of the results in the context of other evidence, and implications for future research. | 10,11 |
| **FUNDING** |  |  |  |
| Funding | 27 | Describe sources of funding for the systematic review and other support (e.g., supply of data); role of funders for the systematic review. | NA |

From: Moher D, Liberati A, Tetzlaff J, Altman DG, The PRISMA Group (2009). Preferred Reporting Items for Systematic Reviews and Meta-Analyses: The PRISMA Statement. PLoS Med 6(7): e1000097. doi:10.1371/journal.pmed1000097
For more information, visit: www.prisma-statement.org.

## Supplemental Material

**Table. S1.** Studies included in the meta-analysis

| Study | Journal | Study type | Date | Country |
| --- | --- | --- | --- | --- |
| **Siegel et al** | American Roentgen Ray Society | Case series | 3-Apr-20 | USA |
| **Pazgan-simon et al** | Polish Archives of Internal Medicine | Case report | 3-Apr-20 | Poland |
| **Yang, X et al** | Clinics and Research in Hepatology and Gastroenterology | Case report | 15-Apr-20 | China |
| **Zhang, J et al** | Allergy (John Wiley and Sons Ltd.) | Retrospective cohort study | 18-Feb-20 | China |
| **Azwar et al** | Indones J Intern Med | Case report | Jan-20 | Indonesia |
| **Jin, X et al** | BMJ | Retrospective cohort study | 17-Mar-20 | China |
| **Nobel et al** | Gastroenterology | Case-Control Study | 8-Apr-20 | USA |
| **Lin et al** | BMJ | Retrospective cohort study | 24-Mar-20 | China |
| **Zhou et al** | Gastroenterology | Retrospective cohort study | 12-Mar-20 | China |
| **Pan et al** | The American Journal of GASTROENTEROLOGY | Cross-sectional study | 14-Apr-20 | China |
| **Poggiali et al** | European Journal of Case Reports in Internal Medicine | Case series | 26-Mar-20 | Italy |
| **Cholankeril et al** | Gastroenterology | Retrospective cohort study | 4-Apr-20 | USA |
| **Fu et al** | Digestive Diseases and Sciences (Springer) | Case report | 28-Apr-20 | China |
| **Li et al** | World Journal of Clinical Cases | Case report | 6-Apr-20 | China |
| **Saeed et al** | British Journal of Surgery | Retrospective cohort study | 28-Apr-20 | Norway |
| **Arashiro et al.** | Journal of travel medicine | case-report | 16-Apr-20 | Japan |
| **Yang F et al** | Journal Of Medical Virology | Retrospective Cohort Study | 15-Apr-20 | China |
| **Guillen, E et al** | American Journal of Transplantation | Case Report | 20-Mar-20 | Spain |
| **Chen, Y et al** | Journal Of Medical Virology | Retrospective Cohort Study | 3-Apr-20 | China |
| **Chen Q et al** | Journal of Medical Virology | SHORT COMMUNICATION | 12-Mar-20 | China |
| **Huang et al** | The Lancet | Prospective Cohort study | 15-Feb-20 | China |
| **Chen, N et al** | The Lancet | Retrospective Cohort Study | 15-Feb-20 | China |
| **Wang et al** | JAMA | Case series | 17-Mar-20 | China |
| **Xu, X et al** | BMJ | retrospective case series | 13-Feb-20 | China |
| **Fan et al** | MedRxiv | Retrospective Cohort Study | 28-Feb-20 | China |
| **Zhang, B et al** | MedRxiv | Retrospective Cohort Study | 27-Feb-20 | China |
| **Huang Y et al** | MedRxiv | Retrospective Cohort Study | 5-Mar-20 | China |
| **Wan, S et al** | Journal of Medical Virology | Retrospective Cohort Study | 6-Mar-20 | China |
| **Zhang, Y et al** | Liver International | Retrospective Cohort Study | 30-Mar-20 | China |
| **Xu, Z et al** | The Lancet | Case Report | 17-Feb-20 | China |
| **Arentz et al** | JAMA | Retrospective Cohort Study | 17-Mar-20 | USA |
| **Hajifathalian et al** | Gastroenterology | Retrospective Cohort Study | 1-May-20 | USA |
| **Kujawski et al** | MedRxiv | Retrospective Cohort Study | 12-Mar-20 | USA |
| **Young et al** | JAMA | Case series | 3-Mar-20 | Singapore |
| **Sun et al** | Clinical Infectious Diseases | retrospective case-control | 25-Mar-20 | Singapore |
| **Pung et al** | The Lancet | Retrospective Cohort Study | 16-Mar-20 | Singapore |
| **Tabata et al** | MedRxiv | Retrospective Cohort Study | 7-Apr-20 | Japan |
| **Kluytmans et al** | MedRxiv | cross-sectional study | 31-Mar-20 | Netherlands |
| **Qian et al** | Quarterly Journal of Medicine | Retrospective case series | 17-Mar-20 | China |
| **Luo et al** | Clinical Gastroenterology and Hepatology | Retrospective cohort study | 20-Mar-20 | China |
| **Zhou F et al** | Lancet | Retrospective Cohort Study | 11-mar-20 | China |
| **Chen T et al** | BMJ | Retrospective Case Series | 26-Mar-20 | China |
| **Xu H et al** | MedRxiv | Retrospective Cohort Study | 30-Mar-20 | China |
| **Shi S et al** | JAMACardio | Retrospective Cohort Study | 25-Mar-20 | China |
| **Han R et al** | Lancet | Retrospective Cohort Study | 12-Jun-20 | China |
| **Xu S et al** | MedRxiv | Retrospective analysis | pre-print | China |
| **Ma L et al** | MedRxiv | Retrospective Study | 30-MAr-20 | China |
| **Liu L et al** | Microbes and infection | Retrospective Study | 18-May-20 | China |
| **Mao L et al** | JAMANEurology | Case Series | 10-Apr-20 | China |
| **Ai JW et al** | Frontiers in Medicine | Cross sectional study | 9-Jun-20 | China |
| **Liu Y et al** | MedRxiv | Retrospective Study | preprint | China |
| **Shu L et al** | Lancet | Retrospective Cohort Study | preprint | China |
| **Wei L et al** |  |  |  | China |
| **Zhao Z et al** | MedRxiv | Retrospective Study | 6-Mar-20 | China |
| **Zhao W et al** | MedRxiv | Retrospective Cohort Study | 30-Mar-20 | China |
| **Yang P et al** | MedRxiv | Retrospective Study | 3-Mar-20 | China |
| **Li K et al** | Investigative Radiology | Retrospective Study | 1-June-20 | China |
| **Qi D et al** | MedRxiv | Retrospective Descriptive study | 3-Mar-20 | China |
| **Wen Y et al** | MedRxiv | Retrospective Study | 23-Mar-20 | China |
| **Xu Y et al** | MedRxiv | Retrospective Observational Study | 6-Mar-20 | China |
| **Yan S et al** | MedRxiv | Retrospective Study | 23-Mar-20 | China |
| **Wang L et al** | European Respiratory Journal | Retrospective Study | 23-Apr-20 | China |
| **Chen X et al** | MedRxiv | Observational study | 6-Mar-20 | China |
| **Liu S et al** | BMC Infectious Diseases | Cohort Study | 6-Jun-20 | China |
| **Yao et al** | Chinese Journal of Hepatology | Retrospective Study | 20-Mar-20 | China |
| **Tian S et al** | MedRxiv | Retrospective study | 23-Mar-20 | China |
| **Lu H et al** | MedRxiv | Descriptive Study | 23-Feb-20 | China |
| **Fu H et al** | MedRxiv | Observational Study | 23-Mar-20 | China |
| **Fu H et al** | MedRxiv | Observational Study | 1-Mar-2020 | China |
| **Chen D et al** | MedRxiv | Retrospective Study | 29-Feb-20 | China |
| **Kuang et al** |  |  |  | China |
| **Rubin et al** | Journal of Clinical And Translational Science | Cohort | 7-Jun-20 | USA |
| **Covid-19 National Emergency Response Center South Korea** | Osong Public Health and Research Perspectives | Case Series | 11-Feb-20 | South Korea |
| **Pung et al** | Lancet | Retrospective | 17-Mar-20 | Singapore |
| **Wolfel et al** | Nature | Case series | 1-Apr-20 | Germany |
| **Dreher et al** | Dtsch Arztebl International | Retrospective | Apr-20 | Germany |
| **Gritti et al** | MedRxiv | Observational Cohort Study | Jun-20 | Italy |
| **Spiteri et al** | Euro Surveillance | Surveillance | 5-Mar-20 | Germany, Finland, Italy, Russia, Spain, France, Sweden, and Belgium |
| **Covid-19 National Incident Room Surveillance Team Australia** | Communicable Diseases Intelligence (2018) | Epidemiology Report | Mar-20 | Australia |
| **An P et. al.** |  | Case series | Jan-20 | China |
| **Chan et. al.** |  | Case series | Jan-20 | China |
| **Chang et al** |  | Case series | Jan-20 | China |
| **Chen et al** |  | Case series | Jan-20 | China |
| **Cheung et al** |  | Case series | Feb-20 | China |
| **Fan H et al** |  | Case series | 16-Feb-20 | China |
| **Fernandez-Ruiz et al** |  | Case series | Mar-20 | Spain |
| **Guan et al** |  | Retrospective cohort | 29-Jan-20 | China |
| **Han et al** |  | Retrospective cohort | Feb-20 | China |
| **Hsih et al** |  | Case series | 19-Feb-20 | Taiwan |
| **Huang et al** |  | Case series | 1-Feb-20 | China |
| **Huang et al** |  | Case series | - | China |
| **Kim ES et al** |  | Case series | 17-Feb-20 | Korea |
| **Klopfenstein et al** |  | Case series | 17-Mar-20 | France |
| **Liu K et al** |  | Case series | 24-Jan-20 | China |
| **Lechien et al** |  | Case series | - | Europe |
| **Liu Y et al** |  | Case series | Jan-20 | China |
| **Pan F et al** |  | Retrospective cohort | 2-feb-20 | China |
| **Redd et al** |  | Retrospective cohort | 2-Apr-20 | US |
| **Ren et al** |  | Case series | 29-Dec-19 | China |
| **Shi H et al** |  | Retrospective cohort | 23-Jan-20 | China |
| **Song et al** |  | Case series | 27-Jan-20 | China |
| **Wan Y et al** |  | Case series | 6-Mar-20 | China |
| **Wang L (b) et. al.** |  | Case series | 12-Feb-20 | China |
| **Wang L (c) et. al.** |  | Case series | 6-Feb-20 | China |
| **Wang X et. al.** |  | Case series | 12-Feb-20 | China |
| **Wang Z et. al.** |  | Case series | 24-Jan-20 | China |
| **Wei X-S et. al.** |  | Case series | 7-Feb-20 | China |
| **Wu J (a) et. al.** |  | Retrospective cohort | 14-Feb-20 | China |
| **Wu J (b) et al** |  | Case series | Feb-20 | China |
| **Wu Y et. al.** |  | Case series | 15-Mar-20 | China |
| **Xia X et. al.** |  | Case series | - | China |
| **Xiao F et. al.** |  | Case series | 14-Feb-20 | China |
| **Xie H et. al.** |  | Case series | 23-Feb-20 | China |
| **Xiong Y et. al.** |  | Case series | 5-Feb-20 | China |
| **Xu X et al** |  | Case series | 4-Feb-20 | China |
| **Yu P et. al.** |  | Case series | 23-Jan-20 | China |
| **Zhang J (b) et. al** |  | Case series | 10-Feb-20 | China |
| **Zhao X-Y et. al.** |  | Case series | 10-Feb-20 | China |
| **Zhou S et. al.** |  | Case series | 30-Jan-20 | China |
| **Zou L et. al.** |  | Case series | 26-Jan-20 | China |

**Table. S2.** The Methodical Index for Non-randomized Studies (MINORS)

| Study | 1 | 2 | 3 | 4 | 5 | 6 | 7 | 8 | Total |
| --- | --- | --- | --- | --- | --- | --- | --- | --- | --- |
| Siegel et al | 1 | 1 | 0 | 0 | 0 | 0 | 0 | 0 | 2 |
| Pazgan-simon et al | 1 | 0 | 0 | 1 | 0 | 0 | 0 | 0 | 2 |
| Yang, X et al | 1 | 0 | 0 | 1 | 0 | 0 | 0 | 0 | 2 |
| Zhang, J et al | 2 | 2 | 2 | 1 | 0 | 0 | 0 | 1 | 8 |
| Azwar et al | 1 | 1 | 0 | 1 | 0 | 1 | 0 | 0 | 4 |
| Jin, X et al | 2 | 2 | 2 | 2 | 0 | 1 | 1 | 1 | 11 |
| Nobel et al | 2 | 2 | 2 | 1 | 0 | 0 | 0 | 1 | 8 |
| Lin et al | 1 | 1 | 0 | 2 | 0 | 1 | 0 | 1 | 6 |
| Zhou et al | 2 | 1 | 0 | 1 | 0 | 0 | 0 | 1 | 5 |
| Pan et al | 2 | 1 | 1 | 0 | 0 | 0 | 0 | 1 | 5 |
| Poggiali et al | 2 | 0 | 0 | 1 | 0 | 1 | 0 | 2 | 6 |
| Cholankeril et al | 2 | 1 | 2 | 0 | 0 | 0 | 0 | 0 | 5 |
| Fu et al | 2 | 0 | 1 | 1 | 0 | 0 | 0 | 0 | 4 |
| Li et al | 2 | 0 | 2 | 0 | 0 | 0 | 0 | 0 | 4 |
| Saeed et al | 2 | 1 | 0 | 1 | 0 | 0 | 0 | 1 | 5 |
| Arashiro et al. | 2 | 0 | 0 | 2 | 0 | 0 | 0 | 1 | 5 |
| Yang F et al | 1 | 0 | 2 | 0 | 0 | 0 | 0 | 1 | 4 |
| Guillen, E et al | 2 | 1 | 1 | 1 | 0 | 1 | 0 | 2 | 8 |
| Chen, Y et al | 1 | 1 | 0 | 2 | 0 | 1 | 0 | 1 | 6 |
| Chen Q et al | 2 | 1 | 0 | 1 | 0 | 0 | 0 | 1 | 5 |
| Huang et al | 2 | 1 | 1 | 0 | 0 | 0 | 0 | 1 | 5 |
| Chen, N et al | 2 | 0 | 1 | 1 | 0 | 0 | 0 | 2 | 6 |
| Wang et al | 2 | 0 | 2 | 0 | 0 | 0 | 0 | 2 | 6 |
| Xu, X et al | 2 | 1 | 0 | 1 | 0 | 0 | 0 | 2 | 6 |
| Fan et al | 2 | 0 | 1 | 1 | 0 | 0 | 0 | 2 | 6 |
| Zhang, B et al | 2 | 1 | 2 | 0 | 0 | 0 | 0 | 2 | 7 |
| Huang Y et al | 2 | 0 | 1 | 1 | 0 | 0 | 0 | 2 | 6 |
| Wan, S et al | 2 | 0 | 2 | 0 | 0 | 0 | 0 | 2 | 6 |
| Zhang, Y et al | 2 | 1 | 0 | 1 | 0 | 0 | 0 | 2 | 6 |
| Xu, Z et al | 2 | 0 | 0 | 0 | 0 | 0 | 0 | 2 | 4 |
| Arentz et al | 1 | 0 | 2 | 0 | 0 | 0 | 0 | 1 | 4 |
| Hajifathalian et al | 2 | 1 | 1 | 1 | 0 | 0 | 0 | 2 | 7 |
| Kujawski et al | 1 | 1 | 0 | 1 | 0 | 0 | 0 | 1 | 4 |
| Young et al | 2 | 1 | 0 | 1 | 0 | 0 | 0 | 2 | 6 |
| Sun et al | 2 | 0 | 0 | 0 | 0 | 0 | 0 | 2 | 4 |
| Pung et al | 1 | 0 | 2 | 0 | 0 | 0 | 0 | 1 | 4 |
| Tabata et al | 2 | 1 | 0 | 1 | 0 | 0 | 0 | 1 | 5 |
| Kluytmans et al | 2 | 1 | 1 | 0 | 0 | 0 | 0 | 1 | 5 |
| Qian et al | 2 | 0 | 0 | 1 | 0 | 1 | 0 | 2 | 6 |
| Luo et al | 0 | 1 | 1 | 1 | 2 | 3 | 0 | 1 | 9 |
| Zhou F et al | 2 | 0 | 1 | 4 | 0 | 4 | 0 | 2 | 13 |
| Chen T | 1 | 1 | 1 | 2 | 1 | 0 | 0 | 2 | 8 |
| Xu H et al | 2 | 1 | 0 | 3 | 0 | 0 | 0 | 1 | 7 |
| Shi S et al | 2 | 1 | 1 | 0 | 0 | 0 | 0 | 2 | 6 |
| Han R et al | 1 | 0 | 0 | 0 | 0 | 1 | 0 | 2 | 4 |
| Xu S et al | 0 | 1 | 1 | 1 | 2 | 3 | 0 | 1 | 9 |
| Ma L et al | 2 | 0 | 1 | 3 | 0 | 3 | 0 | 1 | 9 |
| Liu L et al | 2 | 0 | 2 | 0 | 0 | 0 | 0 | 2 | 6 |
| Mao L et al | 2 | 1 | 0 | 1 | 0 | 0 | 0 | 2 | 6 |
| Ai JW et al | c | 0 | 0 | 0 | 0 | 0 | 0 | 2 | 4 |
| Shu L et al | 1 | 0 | 2 | 0 | 0 | 0 | 0 | 1 | 4 |
| Wei L et al | 2 | 1 | 1 | 1 | 0 | 0 | 0 | 2 | 7 |
| Zhao Z et al | 1 | 1 | 0 | 1 | 0 | 0 | 0 | 1 | 4 |
| Zhao W et al | 2 | 0 | 2 | 0 | 0 | 0 | 0 | 2 | 6 |
| Yang P et al | 2 | 1 | 0 | 1 | 0 | 0 | 0 | 2 | 6 |
| Li K et al | 2 | 0 | 0 | 0 | 0 | 0 | 0 | 2 | 4 |
| Qi D et al | 2 | 1 | 0 | 1 | 0 | 0 | 0 | 1 | 5 |
| Wen Y et al | 2 | 1 | 1 | 0 | 0 | 0 | 0 | 1 | 5 |
| Xu Y et al | 1 | 0 | 0 | 0 | 0 | 1 | 0 | 2 | 4 |
| Yan S et al | 0 | 1 | 1 | 1 | 2 | 3 | 0 | 1 | 9 |
| Wang L et al | 2 | 0 | 1 | 3 | 0 | 4 | 0 | 2 | 11 |
| Chen X et al | 2 | 1 | 0 | 3 | 0 | 0 | 0 | 1 | 7 |
| Liu S et al | 2 | 1 | 1 | 0 | 0 | 0 | 0 | 2 | 6 |
| Fan L et al | 1 | 0 | 1 | 0 | 0 | 1 | 0 | 2 | 5 |
| Yao et al | 0 | 1 | 1 | 1 | 2 | 3 | 0 | 1 | 9 |
| Tian S et al | 2 | 0 | 1 | 3 | 0 | 4 | 0 | 1 | 10 |
| Lu H et al | 2 | 1 | 0 | 1 | 0 | 0 | 0 | 2 | 6 |
| Fu H et al | 2 | 1 | 1 | 0 | 0 | 0 | 0 | 1 | 5 |
| Fu H et al | 2 | 1 | 0 | 3 | 0 | 0 | 0 | 1 | 7 |
| Chen D et al | 2 | 1 | 1 | 0 | 0 | 0 | 0 | 2 | 6 |
| Kuang et al | 1 | 0 | 0 | 0 | 0 | 1 | 0 | 2 | 4 |
| Rubin et al | 0 | 1 | 1 | 1 | 2 | 3 | 0 | 1 | 9 |
| COVID-19 National Emergency Response Center | 2 | 0 | 1 | 3 | 0 | 4 | 0 | 2 | 12 |
| Pung et al | 2 | 2 | 0 | 1 | 0 | 0 | 0 | 1 | 6 |
| Wolfel | 2 | 1 | 0 | 1 | 0 | 0 | 0 | 1 | 5 |
| Dreher et al | 2 | 1 | 1 | 0 | 0 | 0 | 0 | 1 | 5 |
| Gritti et al | 2 | 0 | 0 | 1 | 0 | 1 | 0 | 2 | 6 |
| Spiteri et al | 0 | 1 | 1 | 1 | 2 | 3 | 0 | 1 | 9 |
| Covid-19 National Incident Room Surveillance Team Australia | 2 | 0 | 1 | 4 | 0 | 4 | 0 | 2 | 13 |
| An P et. al. | 1 | 1 | 1 | 2 | 1 | 0 | 0 | 2 | 8 |
| Chan F-W et.al. | 2 | 1 | 0 | 3 | 0 | 0 | 0 | 1 | 7 |
| Chang D et. al. | 2 | 1 | 1 | 0 | 0 | 0 | 0 | 2 | 6 |
| Chen Q (b) et. al. | 1 | 0 | 0 | 0 | 0 | 1 | 0 | 2 | 4 |
| Cheung K et. al. | 0 | 1 | 1 | 1 | 2 | 3 | 0 | 1 | 9 |
| Fan H et. al. | 2 | 0 | 1 | 3 | 0 | 3 | 0 | 1 | 9 |
| FernandezRuiz et. al. | 2 | 0 | 2 | 0 | 0 | 0 | 0 | 2 | 6 |
| Guan W-j et. al. | 2 | 1 | 0 | 1 | 0 | 0 | 0 | 2 | 6 |
| Han C et. al. | c | 0 | 0 | 0 | 0 | 0 | 0 | 2 | 4 |
| Hsieh W-H et. al. | 1 | 0 | 2 | 0 | 0 | 0 | 0 | 1 | 4 |
| Huang R et. al. | 2 | 1 | 1 | 1 | 0 | 0 | 0 | 2 | 7 |
| Huang WH et. al. | 1 | 1 | 0 | 1 | 0 | 0 | 0 | 1 | 4 |
| Kim ES et. al. | 2 | 0 | 2 | 0 | 0 | 0 | 0 | 2 | 6 |
| Klopfenstein T et. al. | 2 | 1 | 0 | 1 | 0 | 0 | 0 | 2 | 6 |
| Liu K et. al. | 2 | 0 | 0 | 0 | 0 | 0 | 0 | 2 | 4 |
| Lechien J et. al. | 2 | 1 | 0 | 1 | 0 | 0 | 0 | 1 | 5 |
| Liu Y et. al. | 2 | 1 | 1 | 0 | 0 | 0 | 0 | 1 | 5 |
| Pan F et. al. | 1 | 0 | 0 | 0 | 0 | 1 | 0 | 2 | 4 |
| Redd W et. al. | 0 | 1 | 1 | 1 | 2 | 3 | 0 | 1 | 9 |
| Ren L et. al. | 2 | 0 | 1 | 3 | 0 | 4 | 0 | 2 | 11 |
| Shi H et. al. | 2 | 1 | 0 | 3 | 0 | 0 | 0 | 1 | 7 |
| Song F et. al. | 2 | 1 | 1 | 0 | 0 | 0 | 0 | 2 | 6 |
| Wan Y et. al. | 1 | 0 | 1 | 0 | 0 | 1 | 0 | 2 | 5 |
| Wang L (b) et. | 0 | 1 | 1 | 1 | 2 | 3 | 0 | 1 | 9 |
| Wang L (c) et. al. | 2 | 0 | 1 | 3 | 0 | 4 | 0 | 1 | 10 |
| Wang X et. al. | 2 | 1 | 0 | 1 | 0 | 0 | 0 | 2 | 6 |
| Wang Z et. al. | 2 | 1 | 1 | 0 | 0 | 0 | 0 | 1 | 5 |
| Wei X-S et. al. | 2 | 1 | 0 | 3 | 0 | 0 | 0 | 1 | 7 |
| Wu J (a) et. al. | 2 | 1 | 1 | 0 | 0 | 0 | 0 | 2 | 6 |
| Wu J (b) et al | 1 | 0 | 0 | 0 | 0 | 1 | 0 | 2 | 4 |
| Wu Y et. al. | 0 | 1 | 1 | 1 | 2 | 3 | 0 | 1 | 9 |
| Xia X et. al. | 2 | 0 | 1 | 3 | 0 | 4 | 0 | 2 | 12 |
| Xiao F et. al. | 2 | 2 | 0 | 1 | 0 | 0 | 0 | 1 | 6 |
| Xie H et. al. | 1 | 0 | 0 | 0 | 0 | 1 | 0 | 2 | 4 |
| Xiong Y et. al. | 2 | 1 | 0 | 3 | 0 | 0 | 0 | 1 | 7 |
| Xu X et al | 2 | 1 | 1 | 0 | 0 | 0 | 0 | 2 | 6 |
| Yu P et. al. | 1 | 0 | 0 | 0 | 0 | 1 | 0 | 2 | 4 |
| Zhang J (b) et. al. | 0 | 1 | 1 | 1 | 2 | 3 | 0 | 1 | 9 |
| Zhao X-Y et. al. | 2 | 0 | 1 | 3 | 0 | 4 | 0 | 1 | 10 |
| Zhou S et. al. | 2 | 1 | 0 | 1 | 0 | 0 | 0 | 1 | 5 |
| Zou L et. al. | 2 | 1 | 1 | 0 | 0 | 0 | 0 | 1 | 5 |

**Table. S3.**
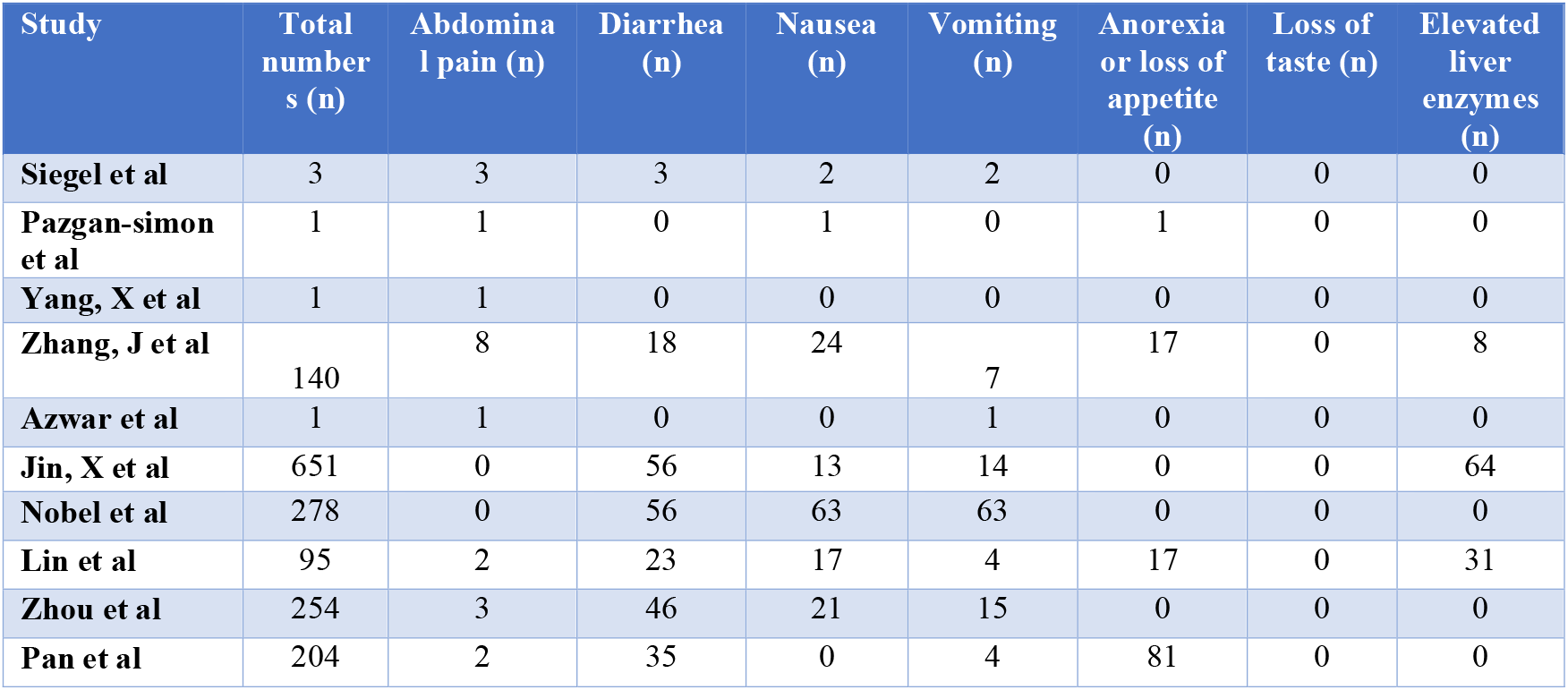
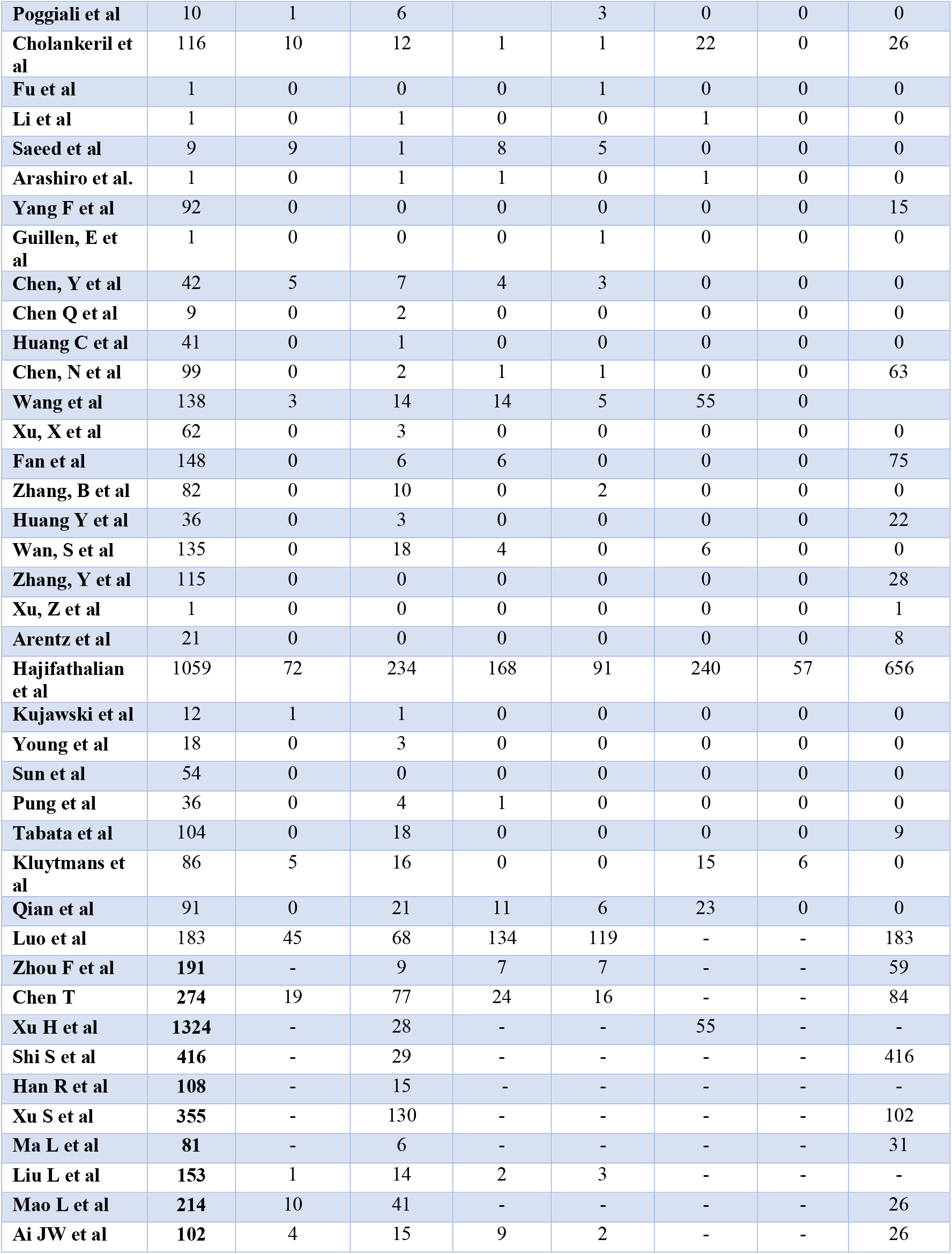
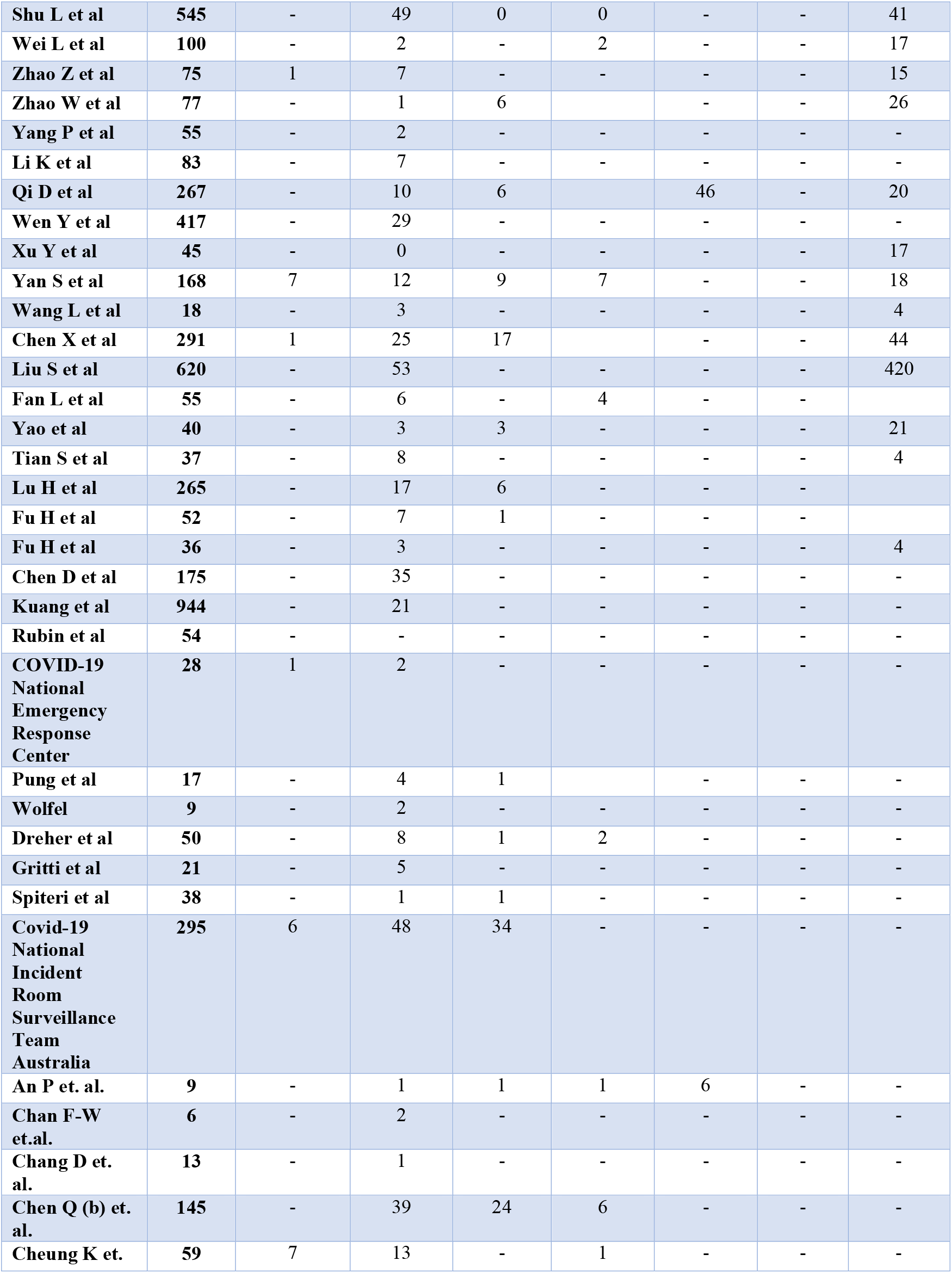
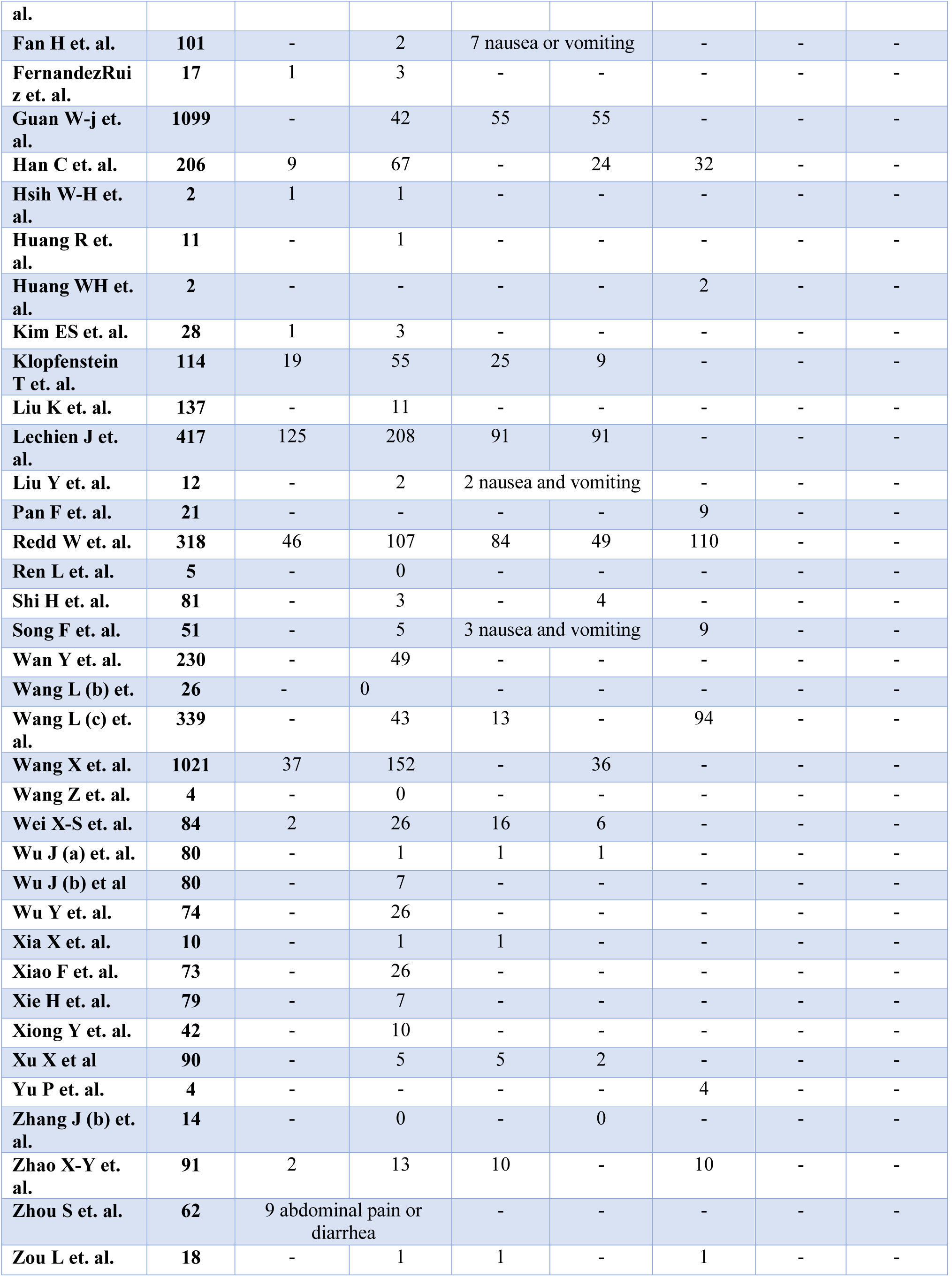
Gastrointestinal symptoms

**Table. S4.** Sensitivity Analysis

| References | No. of patients | Abdominal pain (n) | Diarrhea (n) | Nausea (n) | Vomiting (n) | Anorexia or loss of appetite (n) | Loss of taste (n) | Elevated liver enzymes (n) |
| --- | --- | --- | --- | --- | --- | --- | --- | --- |
| **Zhang, J et al** | 140 | 8 | 18 | 24 | 7 | 17 |  | 8 |
| **Jin, X et al** | 651 | 0 | 56 | 13 | 14 | 0 | 0 | 64 |
| **Lin et al** | 95 | 2 | 23 | 17 | 4 | 17 | 0 | 31 |
| **Zhou et al** | 254 | 3 | 46 | 21 | 15 | 0 | 0 | 0 |
| **Pan et al** | 204 | 2 | 35 | 0 | 4 | 81 | 0 | 0 |
| **Cholankeril et al** | 116 | 10 | 12 | 1 | 1 | 22 | 0 | 26 |
| **Saeed et al** | 9 | 9 | 1 | 8 | 5 | 0 | 0 | 0 |
| **Yang F et al** | 92 | 0 | 0 | 0 | 0 | 0 | 0 | 15 |
| **Chen, Y et al** | 42 | 5 | 7 | 4 | 3 | 0 | 0 | 0 |
| **Huang, C et al** | 41 | 0 | 1 | 0 | 0 | 0 | 0 | 0 |
| **Chen, N et al** | 99 | 0 | 2 | 1 | 1 | 0 | 0 | 63 |
| **Xu, X et al** | 62 | 0 | 3 | 0 | 0 | 0 | 0 | 0 |
| **Fan et al** | 148 | 0 | 6 | 6 | 0 | 0 | 0 | 75 |
| **Zhang, B et al** | 82 | 0 | 10 | 0 | 2 | 0 | 0 | 0 |
| **Huang Y et al** | 36 | 0 | 3 | 0 | 0 | 0 | 0 | 22 |
| **Wan, S et al** | 135 | 0 | 18 | 4 | 0 | 6 | 0 | 0 |
| **Zhang, Y et al** | 115 | 0 | 0 | 0 | 0 | 0 | 0 | 28 |
| **Arentz et al** | 21 | 0 | 0 | 0 | 0 | 0 | 0 | 8 |
| **Hajifathalian et al** | 1059 | 72 | 234 | 168 | 91 | 240 | 57 | 656 |
| **Kujawski et al** | 12 | 1 | 1 | 0 | 0 | 0 | 0 | 0 |
| **Sun et al** | 54 | 0 | 0 | 0 | 0 | 0 | 0 | 0 |
| **Pung et al** | 36 | 0 | 4 | 1 | 0 | 0 | 0 | 0 |
| **Tabata et al** | 104 | 0 | 18 | 0 | 0 | 0 | 0 | 9 |
| **Kluytmans et al** | 86 | 5 | 16 | 0 | 0 | 15 | 6 | 0 |
| **Qian et al** | 91 | 0 | 21 | 11 | 6 | 23 | 0 | 0 |
| **Total** | **3784** | 117 | 535 | 279 | 153 | 421 | 63 | 1005 |

**Table. S5.** Fatality numbers as reported by each study

| **Study** | **Total no. of patients** | **Fatality (n)** | **Country** |
| --- | --- | --- | --- |
| **Siegel et al** | 3 | 0 | USA |
| **Pazgan-simon et al** | 1 | 0 | Poland |
| **Azwar et al** | 1 | 0 | China |
| **Jin, X et al** | 651 | 1 | China |
| **Nobel et al** | 278 | 9 | Indonesia |
| **Lin et al** | 95 | 0 | China |
| **Zhou et al** | 254 | 16 | USA |
| **Pan et al** | 204 | 36 | China |
| **Cholankeril et al** | 116 | 1 | China |
| **Fu et al** | 1 | 0 | China |
| **Li et al** | 1 | 1 | Italy |
| **Saeed et al** | 9 | 1 | USA |
| **Arashiro et al.** | 1 | 1 | China |
| **Yang F et al** | 92 | 92 | China |
| **Chen Q et al** | 9 | 0 | Norway |
| **Huang, C et al** | 41 | 6 | Japan |
| **Chen, N et al** | 99 | 11 | China |
| **Wang et al** | 138 | 6 | Spain |
| **Xu, X et al** | 62 | 0 | China |
| **Fan et al** | 148 | 1 | China |
| **Zhang, B et al** | 82 | 82 | China |
| **Huang Y et al** | 36 | 36 | China |
| **Wan, S et al** | 135 | 1 | China |
| **Zhang, Y et al** | 115 | 1 | China |
| **Xu, Z et al** | 1 | 1 | China |
| **Arentz et al** | 21 | 11 | China |
| **Kujawski et al** | 12 | 0 | China |
| **Young et al** | 18 | 0 | China |
| **Sun et al** | 54 |  | China |
| **Pung et al** | 36 | 0 | China |
| **Qian et al** | 91 | 0 | USA |
| **Luo et al** | 183 | 7 | USA |
| **Zhou F et al** | 191 | 54 | USA |
| **Chen T et al** | 274 | 113 | Singapore |
| **Shi S et al** | 416 | 30 | Singapore |
| **Mao L et al** | 214 | 1 | China |
| **Ai JW et al** | 102 | 3 | China |
| **Liu Y et al** | 109 | 31 | China |
| **Shu L et al** | 545 | 0 | China |
| **Wei et al** | 100 | 3 | China |
| **Zhao W et al** | 77 | 5 | China |
| **Yang P et al** | 55 | 2 | China |
| **Qi D et al** | 267 | 4 | China |
| **Wen Y et al** | 417 | 3 | China |
| **Xu Y et al** | 45 | 0.09 | China |
| **Yan S et al** | 168 | 6 | China |
| **Wang L et al** | 18 | 0 | China |
| **Chen X et al** | 291 | 2 | China |
| **Liu S et al** | 620 | 0 | China |
| **Tian S et al** | 37 | 0 | China |
| **Lu H et al** | 265 | 1 | China |
| **Fu H et al** | 52 | 0 | China |
| **Pung et al** | 17 | 0 | China |
| **Dreher et al** | 50 | 7 | China |
| **Gritti et al** | 21 | 1 | China |
| **Spiteri et al** | 38 | 1 | China |
| **Covid-19**<br>**National**<br>**Incident Room**<br>**Surveillance**<br>**Team**<br>**Australia** | 295 | 3 | China |
| **Fan H et. al.** | 101 | 101 | Italy |
| **FernandezRuiz**<br>**et. al.** | 17 | 5 | Germany,<br>Finland,<br>Italy,<br>Russia,<br>Spain,<br>France,<br>Sweden,<br>and<br>Belgium |
| **Han C et. al.** | 206 | 0 | China |
| **Hsih W-H et.**<br>**al.** | 2 | 0 | China |
| **Huang WH et.**<br>**al.** | 2 | 0 | China |
| **Kim ES et. al.** | 28 | 0 | China |
| **Liu Y et. al.** | 12 | 0 | China |
| **Pan F et. al.** | 21 | 0 | Taiwan |
| **Redd W et. al.** | 318 | 32 | China |
| **Ren L et. al.** | 5 | 1 | China |
| **Shi H et. al.** | 81 | 3 | Korea |
| **Song F et. al.** | 51 | 0 | France |
| **Wan Y et. al.** | 230 | 6 | China |
| **Wang L (b) et.**<br>**al.** | 26 | 0 | Europe |
| **Wang L (c) et.**<br>**al.** | 339 | 4 | China |
| **Wang X et. al.** | 1021 | 0 | China |
| **Wei X-S et. al.** | 84 | 0 | China |
| **Wu Y et. al.** | 74 | 1 | China |
| **Xiong Y et. al.** | 42 | 1 | China |
| **Yu P et. al.** | 4 | 1 | China |
| **Zhao X-Y et. al.** | 91 | 2 | China |
| **Total** | 10,427 | 749 |  |

**Supplementary figure S1.**
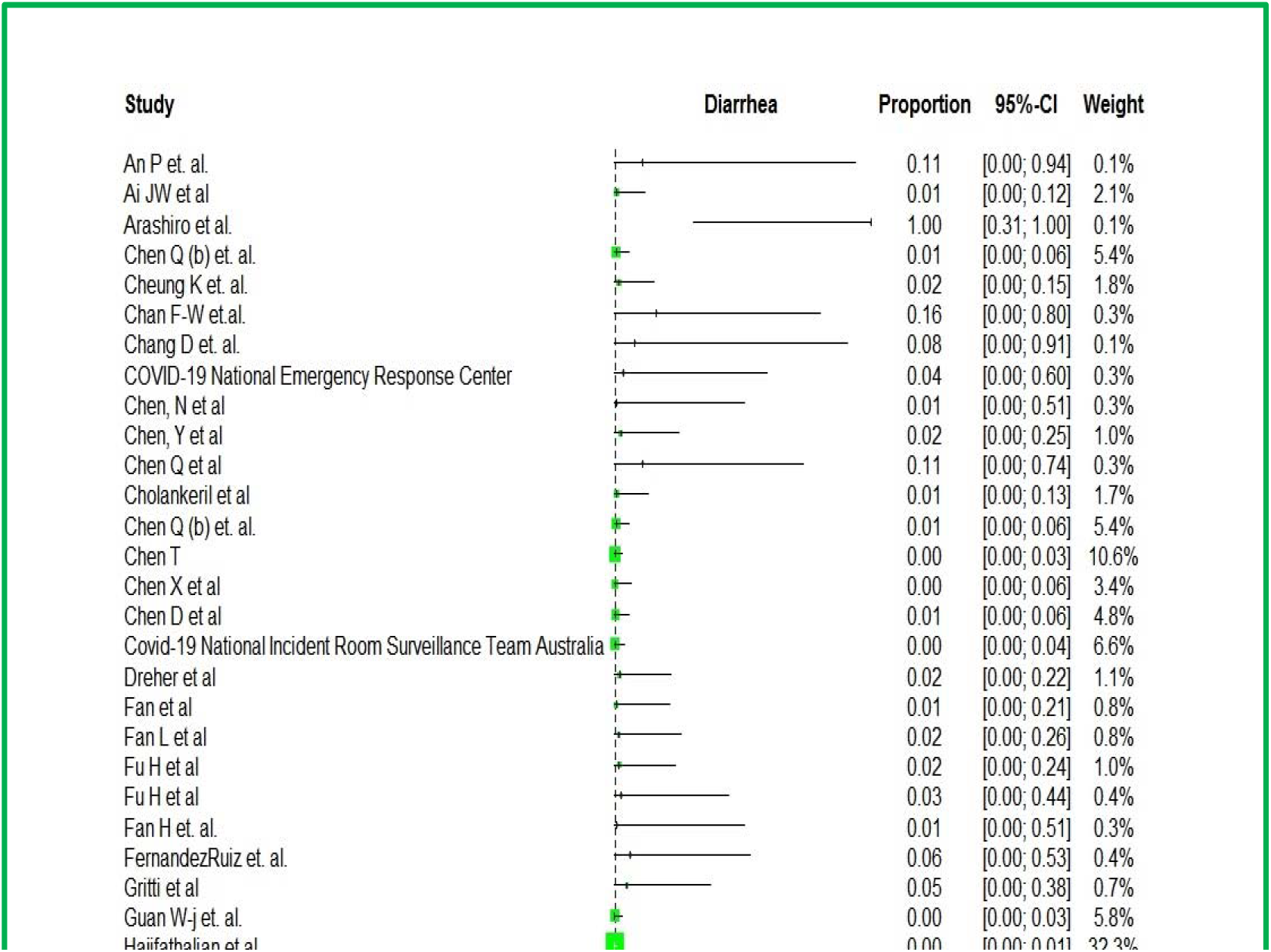
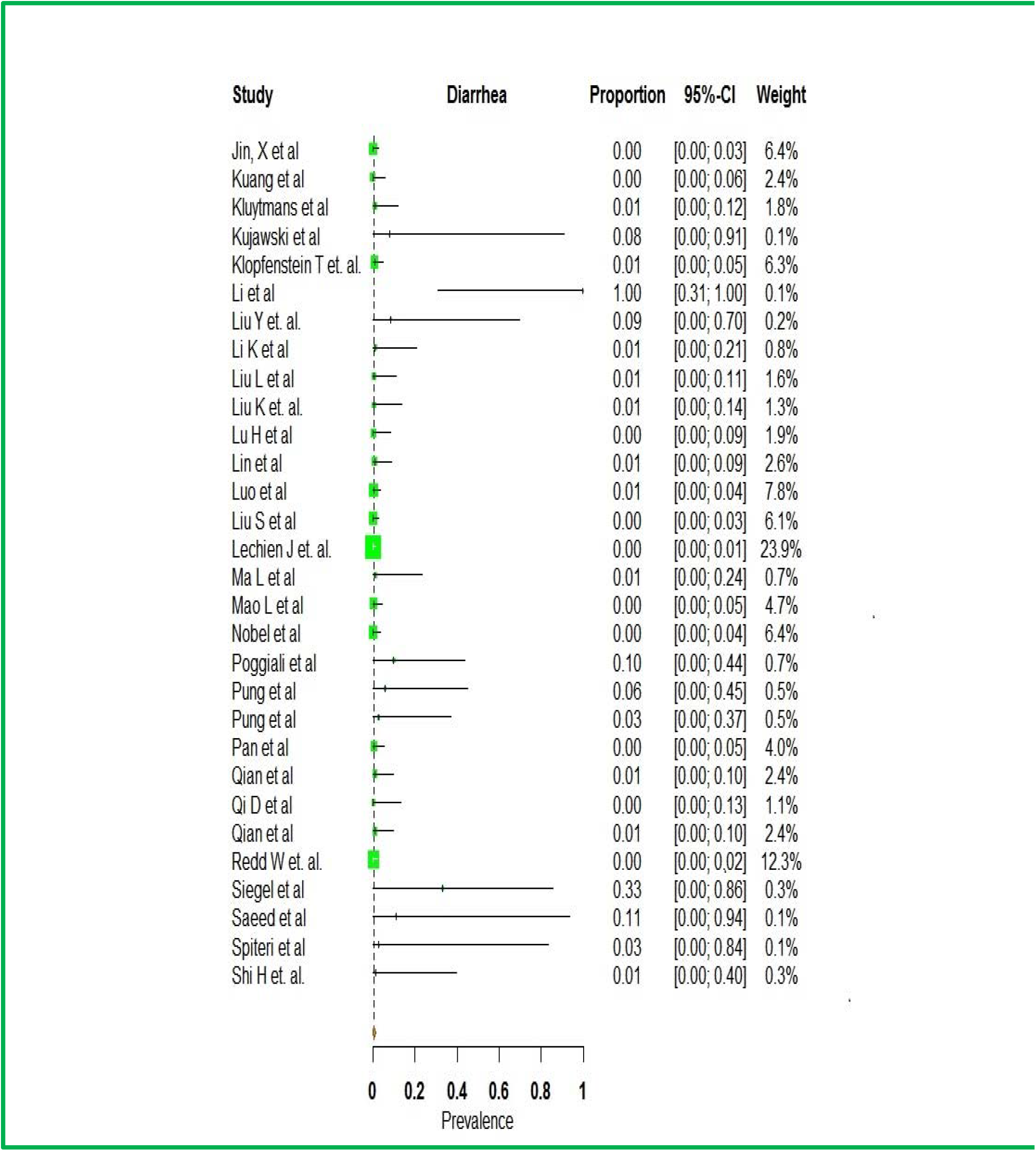
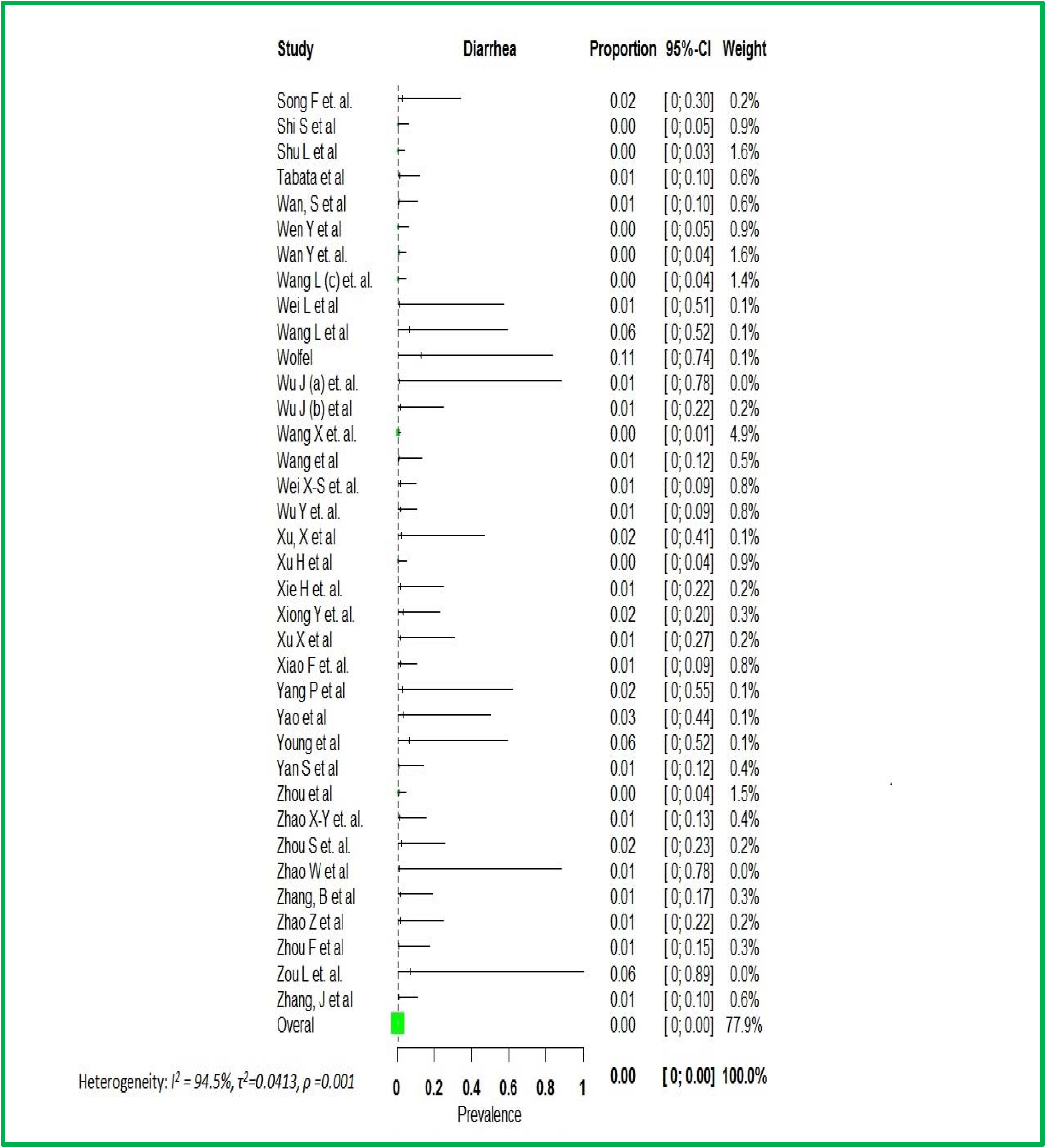
Forest plot depicting prevalence of diarrhea

**Supplementary figure.2.**
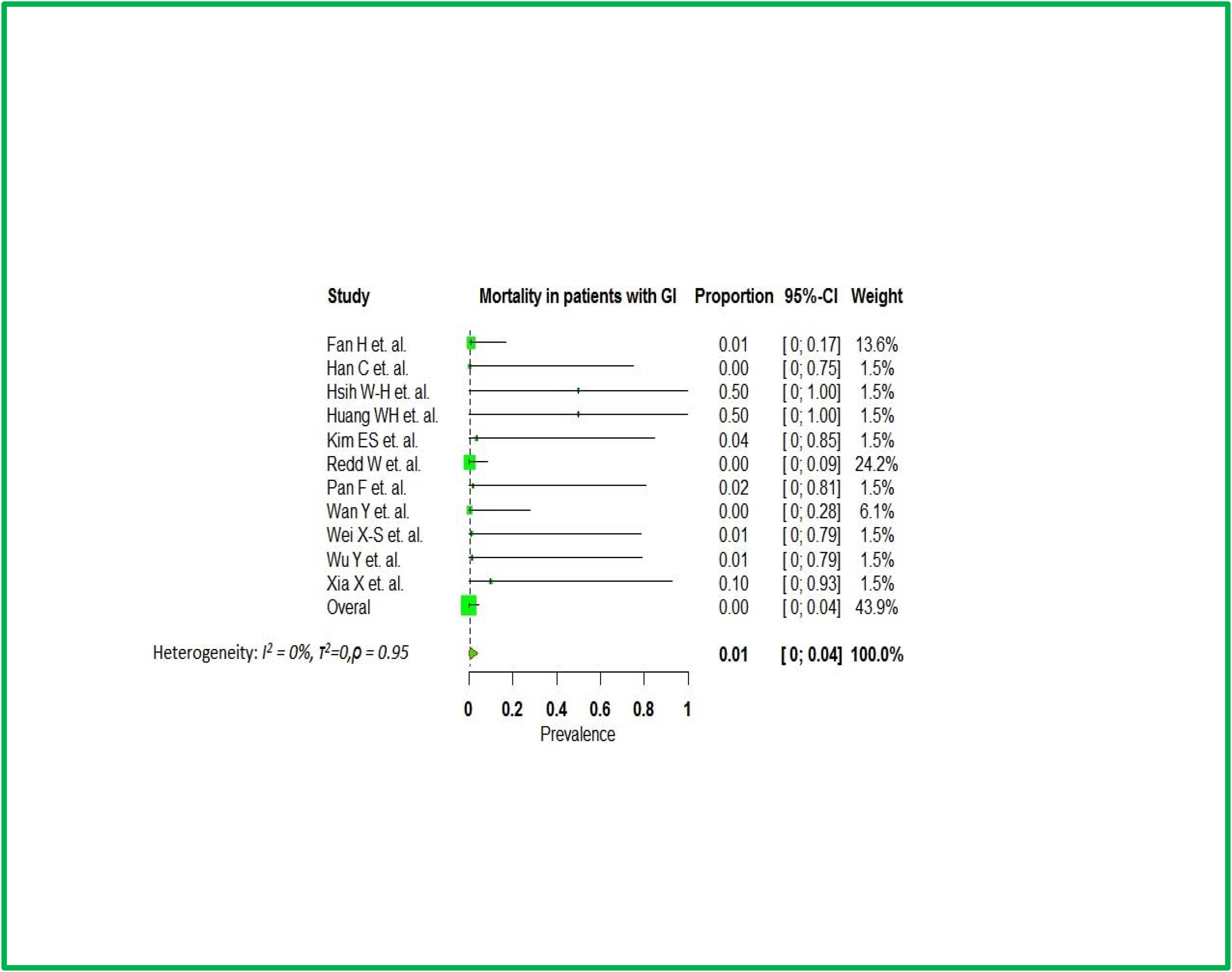

